# Multimodal profiling for prediction of primary resistance to anti–PD-(L)1 therapy in advanced non-small-cell lung cancer: the prospective PIONeeR biomarkers study

**DOI:** 10.64898/2026.01.09.26343779

**Authors:** Fabrice Barlesi, Florence Monville, Frédéric Vely, Laurent Greillier, Florence Sabatier, Jean-Philippe Dales, Anastasiia Bakhmach, Amélie Pouchin, Joseph Ciccolini, Stephane Garcia, Natalie Ngoi, Cyril Foa, Laurent Arnaud, Sivan Bokobza, Andrea Vaglio, Mélanie Karlsen, Paul Dufossé, Clarisse Audigier-Valette, Jacques Letreut, Lilian Laborde, Pierre Milpied, David Pérol, Mohamed Boussena, Celestin Bigarre, Mourad Hamimed, Richard Malkoun, Vanina Leca, Marcellin Landri, Maryannick Le Ray, Marie Roumieux, Julien Mazières, Maurice Pérol, Jacques Fieschi-Meric, Eric Vivier, Sébastien Benzekry

## Abstract

**Background:** Pretreatment prediction of primary resistance to anti–PD-(L)1 therapy in advanced non-small-cell lung cancer (NSCLC) remains an unmet clinical need. Existing biomarkers — including PD-L1 expression and tumour mutational burden (TMB) — are insufficiently discriminatory, and multimodal predictive models targeting primary resistance are still lacking to precisely drive patients treatment strategy.

**Methods:** PIONeeR was a prospective, multicentre biomarker study conducted across 17 centres (NCT03493581). Adults with advanced NSCLC initiating standard-of-care first-line platinum-based chemotherapy plus anti–PD-(L)1 therapy, or second-or-later-line anti– PD-(L)1 monotherapy, were enrolled. Pretreatment multimodal profiling spanned six biological layers: clinical, routine medical biology, high-dimensional circulating immune phenotyping, soluble vascular markers, tumour immune contexture with digital pathology analysis of multiplex immunohistochemistry and immunofluorescence, transcriptomics and genomics. The primary endpoint was prediction of primary resistance (PrR). Thirty-six feature-selection methods and ten machine learning models (360 pipelines) were benchmarked within a .632 optimism-correction framework.

**Results:** Between March 2018 and January 2023, 439 patients were enrolled (269 first-line, 170 subsequent-line); 435 were evaluable for PrR, which occurred in 39.5%. Individual biomarkers showed broad but modest associations with PrR (maximum AUROC 0.64). In the first-line chemo-immunotherapy setting, the multimodal signature achieved an optimism-corrected AUROC of 0.69 ± 0.06 and a Positive Predictive Value (PPV) of 0.51 ± 0.11 against a primary-resistance prevalence of 30%, outperforming PD-L1 (PPV 0.36 ± 0.02) and TMB (PPV 0.35 ± 0.03). In the subsequent-line setting it achieved an AUROC of 0.69 ± 0.08 and a PPV of 0.64 ± 0.08, where PD-L1 alone performed at least as well (PPV 0.68). The 18-feature signature included stromal regulatory T-cell infiltration, circulating transitional B cells, and multiple routine laboratory variables, with treatment-setting-dependent contributions.

**Interpretation:** Despite comprehensive multimodal pretreatment profiling, primary resistance to anti– PD-(L)1 therapy in advanced NSCLC cannot be captured by isolated biomarkers alone. Complete results are publicly accessible through an interactive dashboard (https://compo.inria.fr/pioneer-dashboard/), a reference resource for future meta-analyses. Multimodal integration improved risk stratification while highlighting the strong contribution of routinely available baseline variables. These findings support the development of clinically pragmatic models to identify patients who should access next-generation immunotherapy strategies aimed at overcoming primary resistance to PD-(L)1 inhibition.

## INTRODUCTION

Immune checkpoint inhibitors (ICIs) targeting the PD-1-PD-L1 axis have become a standard component of treatment for advanced wild-type non-small-cell lung cancer (NSCLC) ^1–4^. However, a majority of patients will relapse. Identifying, before treatment initiation, which patients are unlikely to derive meaningful benefit from anti–PD-(L)1-based therapy remains therefore a major unmet need.

Current biomarkers are insufficient for this purpose. PD-L1 expression is clinically useful, but its predictive performance is only moderate, limited to a modest enrichment for prediction of response rather than resistance, and is constrained by assay variability, and spatiotemporal tumour heterogeneity ^2,5–7^. Tumour mutational burden (TMB) has also been investigated extensively, but its performance and clinical utility have been inconsistent across studies and settings ^8–12^. Beyond these markers, multiple candidate predictors of response or resistance have been proposed, including genomic alterations in tumour suppressor genes such as *STK11*, *KEAP1*, and *SMARCA4* ^13–15^, the presence and spatial distribution of tumour-infiltrating lymphocytes and tertiary lymphoid structures ^16,17^, interferon-gamma gene expression signatures^18^, pathomics^19^, and systemic inflammatory indices such as the Lung Immune Prognostic Index (LIPI) ^20^ or circulating tumour DNA^12^. Yet overlap between such markers remains limited, underscoring the difficulty of single-biomarker prediction.

Mechanistically, resistance to anti–PD-(L)1 therapy is likely to reflect the combined influence of tumour-intrinsic, microenvironmental, systemic inflammatory, and host-related factors rather than a single pathway. Multimodal approaches integrating orthogonal biological and clinical inputs may therefore provide a more informative estimate of resistance risk than unimodal biomarkers alone^21–23^. Several such models have now been developed in advanced NSCLC, combining radiomics, pathomics, and genomics^22^, deep learning from tumour histology^19^, or large-scale clinicogenomic data^24,25^, but they share important limitations. Most have used best overall response or overall survival as the primary endpoint — neither of which is interchangeable with primary resistance: response-based endpoints are susceptible to class imbalance and may reward transient benefit, while overall survival is confounded by post-progression therapies and comorbid mortality. Beyond endpoint choice, existing models have rarely integrated the depth of immune phenotyping — spanning tumour-infiltrating immune populations and circulating immune subsets — that the mechanistic landscape most directly motivates, relying instead on radiological, histological, genomic, or routine laboratory inputs. Effective sample sizes for deep multimodal phenotyping have remained modest, and few cohorts have included substantial numbers of patients receiving first-line ICI+CT, now the predominant treatment context in advanced NSCLC. Primary resistance (PrR) — defined according to the Society for Immunotherapy of Cancer (SITC) guidelines^26,27^ — represents a more clinically actionable target; reliable pretreatment predictors of PrR remain underdeveloped.

We therefore designed the prospective multicentre PIONeeR (Precision Immuno-Oncology for advanced Non-Small Cell Lung Cancer Patients with PD1(L1) ICI Resistance) biomarkers study, with an unprecedentedly deep and comprehensive phenotyping strategy. Beyond standard clinical data and routine blood tests, the study integrated systemic immune profiling via high-dimensional flow cytometry and soluble factor quantification, deep characterisation of the tumour immune microenvironment through multiplex immunofluorescence and immunohistochemistry with digital pathology analysis, and —spanning the full biological breadth from circulating host immune state to tumour-intrinsic genomic features. Profiling patients treated with both first-line chemo-immunotherapy and subsequent-line ICI monotherapy (still used in some situations), we asked whether this depth of multimodal integration could unlock meaningful prediction of primary resistance beyond what PD-L1 expression and TMB — the current clinical standards — are able to offer. Through systematic benchmarking of 36 feature-selection methods combined with 10 machine-learning models within a rigorous optimism-correction framework, we derived an 18-feature multimodal signature of primary resistance and characterised its biological interpretation at the patient level.

## MATERIALS AND METHODS

The study was conducted in accordance with the Helsinki declaration, French laws and regulations and the International Conference on Harmonization (ICH) E6 Guideline for Good Clinical Practice. The study was approved by the French ethics committee (Comité de Protection des Personnes Ouest II Angers, no. 2018/08) and the French drug and device regulation agency (Agence Nationale de Sécurité du Médicament, no. 2018020500208). Informed consent was obtained from each participant before any study procedure. The study is registered at ClinicalTrials.gov (NCT03493581).

### Study design

This was a prospective study conducted in 17 centres in France. Eligible patients had histologically confirmed advanced or recurrent NSCLC planned for standard-of-care anti–PD-(L)1-based therapy — first-line platinum-based chemotherapy plus anti–PD-(L)1, or second/third-line anti–PD-(L)1 monotherapy after platinum failure — were anti– PD-(L)1-naive, ≥18 years, ECOG 0–1, with available archival FFPE tumour tissue and adequate organ function. Investigator-assessed radiographic tumour assessment was performed every 6 weeks using RECIST v1.1^28^ (Supplementary Figure 1). Detailed eligibility and follow-up procedures are provided in Supplementary Methods.

### Endpoints

The primary endpoint was PrR, defined according to the 2023 SITC recommendations^26,27^ and adapted to treatment setting: for monotherapy, RECIST v1.1^28^ best overall response of progressive disease or stable disease lasting <6 months from the first ICI dose; for combination therapy, progression within 6 months from the first ICI dose irrespective of best overall response. Secondary endpoints were PFS, OS, objective response, early progression (<3 months) and long-lasting response (absence of progression at 12 months). Detailed evaluability rules are provided in Supplementary Methods.

### Biomarker collection and profiling

#### Clinical data and vital signs

Standard clinical variables including patient and tumour characteristics, ECOG performance status, and baseline vital signs were collected within two weeks before ICI initiation (Supplementary Table 1). Treatment line was retained as a covariate throughout all analyses.

#### Routine haematology and biochemistry

Forty-nine routine laboratory variables, spanning complete blood count with differential and serum biochemistry, were obtained from accredited hospital laboratories within two weeks before ICI initiation. Derived inflammatory indices (neutrophil-to-lymphocyte and monocyte-to-lymphocyte ratios) were computed.

#### Circulating immune profiling

Peripheral blood was collected at baseline for immune-cell phenotyping by flow cytometry on whole blood and PBMCs, covering T-cell, B-cell, NK, monocyte, neutrophil, dendritic-cell and MDSC populations and their activation, exhaustion and checkpoint states. Soluble mediators were quantified by ELISA (sCD25) and cytometric bead array (IFN-γ, TNF, IL-6, IL-10), contributing 142 features in total (137 circulating immune-cell parameters and five soluble mediators; Figure 1C). (Supplementary Methods, Supplementary Table 1).

**Figure 1:**
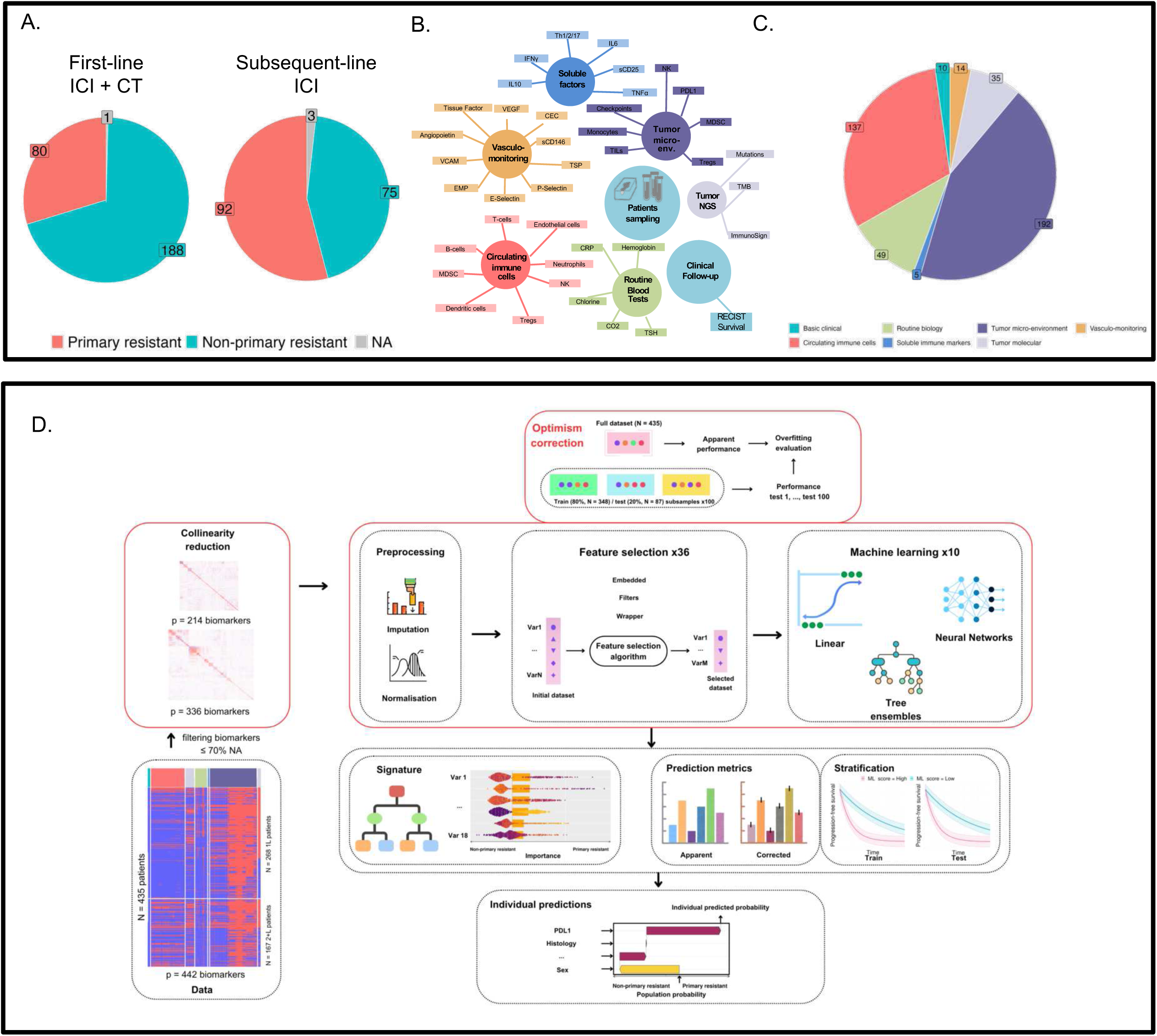
Overview of the multimodal biomarker dataset and analytical workflow. A. Primary endpoint composition by treatment setting. Pie charts show the proportion of patients classified as primary resistant (salmon), non-primary resistant (teal), or non-evaluable (grey) in the first-line ICI + chemotherapy cohort (left; n=269) and the second-or-later-line ICI monotherapy cohort (right; n=170). B. Network representation of the 442 pre-treatment biomarkers collected in the PIONeeR study, grouped by assessment platform: tumour microenvironment assays (including digital pathology-assessed multiplex immunohistochemistry and immunofluorescence) and whole-exome sequencing), circulating immune cell phenotyping (flow cytometry), vasculo-monitoring (soluble endothelial markers), routine laboratory blood tests, and basic clinical parameters. Edges connect biomarkers to their parent platform. NGS: Next-generation sequencing. ImmunoSign: immune contexture transcriptomic signatures^31^. C. Distribution of the 442 pre-treatment biomarkers across seven assessment modalities (basic clinical, n=10; routine blood tests, n=49; tumour microenvironment, n=192; tumour molecular, n=35; circulating immune cells, n=137; soluble immune markers, n=5; vasculo-monitoring, n=14), illustrating the multimodal depth of the dataset. D. End-to-end analytical pipeline. Starting from 442 biomarkers in 439 patients, markers with >70% missing values were removed (336 retained) and iterative variance inflation factor (VIF) analysis was applied to reduce multicollinearity (214 decorrelated features retained; correlation heatmap shown at left). Within an .632 optimism-correction framework using resampling across 100 training/test splits, 36 feature selection algorithms and 10 machine learning models (360 combinations) were benchmarked. Outputs include the selected biomarker signature (shown as a decision tree and ranked SHAP importance plot), optimism-corrected versus apparent prediction metrics, progression-free survival stratification on training and test sets, and individual-level SHAP-based risk explanations. ICI, immune checkpoint inhibitor; CT, chemotherapy; VIF, variance inflation factor; SHAP, SHapley Additive exPlanations; NA, not evaluable; NGS, next-generation sequencing; MDSC, myeloid-derived suppressor cells; VCAM, vascular cell adhesion molecule; ICAM, intercellular adhesion molecule.

#### Circulating endothelial biomarkers

Circulating endothelial cells were isolated from platelet-free plasma using CD146-coated immunomagnetic beads and enumerated by fluorescence microscopy. Soluble adhesion molecules (sVCAM-1, sICAM-1, sE-selectin, sP-selectin) and angiogenic factors were measured by Luminex/ELISA. Endothelial extracellular vesicles were identified by flow cytometry, with tissue-factor activity assessed by an FXa-generation assay^29^.

#### Tumour microenvironment and genomic characterisation

##### Multiplex immunohistochemistry

Up to 22 sequential 4-µm FFPE sections per archival block were processed. PD-L1 expression was assessed by manual tumour proportion score (TPS), visually estimated by three trained pathologists as the percentage of viable tumour cells with partial or complete membranous staining, then either kept as continuous variable or binarized at either a 1% or 50% cut-off. PD-L1 expression also was assessed using the Immunoscore IC kit (clone HDX3, Veracyte). Multiplex IHC panels targeting T-cell (CD3, CD8, PD-1, LAG-3, TIM-3) and myeloid (CD11b, CD14, CD15, HLA-DR, S100A9, LOX-1) markers, and dual CD8-PD-L1 and CD4-FOXP3 staining, were quantified using HALO software within three spatial compartments — whole tumour, stroma, and parenchyma — enabling compartment-specific characterisation. Platform and detection-system details are provided in Supplementary Methods.

##### Multiplex immunofluorescence

Three Opal 6-Plex panels targeting innate immunity, NK/T cells, and B cells were applied to sequential FFPE sections on the Bond RX platform, imaged on a PhenoImager HT, and analysed in HALO Highplex FL after spectral unmixing in inForm.

##### Genomic and transcriptomic sequencing

Tumour DNA and RNA were co-extracted from macrodissected FFPE specimens; matched germline DNA was extracted from peripheral blood. Whole-exome sequencing used the xGen Exome Hyb Panel (IDT) on Illumina NovaSeq, achieving mean coverage of 100× (tumour) and 70× (normal). TMB was calculated from coding non-synonymous variants per megabase^12,30^. Mutational analyses were, selected to match the targeted sequencing panel routinely used by the pathology department. Whole-transcriptome libraries were prepared using QuantSeq 3’ mRNA-Seq (Lexogen) and sequenced on Illumina NextSeq 2000. Two pre-defined immune gene-expression signatures were derived from these data: ImmunoSign21 (21 genes) and ImmunoSign15 (15 genes), profiling T-cell, effector, immune checkpoint, chemokine and interferon-related programmes, as previously described^31^.

### Integrative biostatistics and machine learning

#### Data infrastructure and reproducibility

Analyses were performed in a reproducible containerised environment using three internally developed open-source packages: compo.EDA for biostatistical analyses^32^, compo.tidyML for machine learning analyses^33^, and ROOFS for robust feature selection^34^. A companion website (https://compo.inria.fr/pioneer-website/) was developed, enabling interactive interrogation of biomarker distributions, associations with multiple outcomes, and comprehensive statistical and machine-learning reports.

#### Preprocessing and multicollinearity reduction

Variables with markedly skewed distributions were log-transformed (239 of 442; Supplementary Table 1) and those with ≥70% missingness excluded (106 of 442). Numeric variables were centred and scaled, missing values imputed with median (numeric) or mode (categorical), and multi-level categorical variables one-hot encoded; all preprocessing was embedded within modelling pipelines to prevent information leakage. Iterative Variance Inflation Factor analysis (VIF threshold 5)^35^ yielded a final decorrelated dataset of 214 features.

#### Univariable biostatistical analysis

Associations with binary endpoints were assessed by logistic regression with appropriate parametric or non-parametric tests by sample size (Supplementary Methods); AUROC was computed for each marker. Associations with PFS and OS were assessed by Cox proportional-hazards regression with log-rank testing on Kaplan-Meier curves stratified by maximally selected rank-statistic cutpoints (surv_cutpoint, minprop = 0.1), with C-indices reported. All models were adjusted for PD-L1 TPS (pathologist continuous assessment). Benjamini-Hochberg (BH) false discovery rate (FDR) correction was applied across all biomarkers^36^.

#### Optimism-correction evaluation for machine learning analyses

To obtain unbiased estimates of predictive performance, machine learning modelling pipelines — preprocessing, feature selection, model selection, and hyperparameter tuning — were implemented within a .632 optimism-correction framework^37,38^ using a python package termed *ROOFS* (RObust biOmarker Feature Selection)^34^. Briefly, identical pipelines were applied to the full dataset, yielding apparent performance, and independently to 100 subsamples of 80% of patients, with performance evaluated on the left-out (out-of-bag) patients. The final metric is a weighted average of apparent and test performance^37,38^; full derivation is in Supplementary Methods. Primary metrics were the optimism-corrected AUROC and PPV; secondary metrics were balanced accuracy, sensitivity, NPV and specificity. Calibration was assessed by plotting mean predicted probabilities against observed event rates. Clinical utility was evaluated by decision curve analysis, comparing the net benefit of the final model against PD-L1 expression alone.

Sample size was fixed by the parent clinical study; no prospective power calculation for the prediction model was performed. The 172 primary-resistance events observed among 435 evaluable patients correspond to 0.80 events per candidate predictor across the 214 decorrelated features entering feature selection, and to 9.1 events per variable for the 19-variable final model. This study is reported in accordance with the TRIPOD+AI statement; the completed checklist is provided as a Supplementary file.

#### Feature selection benchmarking

Thirty-six feature-selection methods were benchmarked for both predictive performance and stability across subsamples^39^, spanning embedded methods (LASSO and its variants, Stabl, RENT), filter methods (mutual-information-based scores, ReliefF, mRMR, F/Gini/t-score, hierarchical clustering on the correlation matrix) and wrapper methods (recursive elimination, Shapicant), complemented by univariable PFS-Cox or PrR-logistic filtering with BH-adjusted q < 0.01. All methods were unified within *ROOFS*^34^. The full enumeration with method-specific citations and the stability formula are provided in Supplementary Methods.

#### Predictive modelling

Ten machine-learning models — including logistic regression, random forests, (extreme) gradient boosting, and multi-layer perceptrons — were evaluated within ROOFS^34^ using the scikit-learn library^40^. For tree-based models, maximum tree depth was set to 2, minimum patients per leaf to 40, and number of estimators to 200; logistic regression was fitted without regularisation. Hyperparameter tuning yielded negligible gains and was not retained.

#### PFS stratification analysis

The final model was evaluated for its ability to stratify PFS using a 2/3–1/3 train-test split stratified by k-means clusters (k = 5) on known prognostic variables (PD-L1 at the 1% threshold, histology, performance status, sex). The PrR probability cutpoint was defined in the training set using surv_cutpoint (minprop = 0.1) and propagated to the test set.

#### Explainability analyses

SHapley Additive exPlanation (SHAP) analyses were conducted in both feature and patient dimensions using the kernelshap^41^ and shapviz^42,43^ R packages, with the model trained on the training set and SHAP contributions generated in the test set. Decision trees on signature variables were built using DecisionTreeClassifier (max_depth = 4, min_samples_leaf = 10, ccp_alpha = 0.01) from scikit-learn^40^.

## RESULTS

### Patient clinical characteristics

Between March 2018 and January 2023, 498 patients were prospectively screened, and 439 met inclusion criteria; 435 were evaluable for PrR (Table 1, Supplementary Figure 1). Of these, 268 received first-line chemo-immunotherapy and 167 received second- or third-line anti–PD-(L)1 monotherapy. Median PFS was 10.1 months (95% CI 9.0–14.3) in first-line patients and 3.77 months (2.77–5.43) in subsequent-line patients after median follow-up of 16.9 and 25.5 months, respectively (Supplementary Figure 2). Investigator-assessed overall response rate was 48.8% overall (62.7% chemo-immunotherapy; 25.2% monotherapy). PrR occurred in 39.5% of evaluable patients overall, including 29.9% (80 of 268) of first-line patients and 55.1% (92 of 167) of subsequent-line patients (Figure 1A).

**Table 1:** Patient characteristics. Clinical characteristics of 439 patients enrolled in the PIONeeR study, stratified by treatment setting: first-line platinum-based chemotherapy plus anti–PD-(L)1 therapy (1L CT+ICI, n=269) and second-or-later-line anti–PD-(L)1 monotherapy (2L+ ICI, n=170). Percentages are calculated on non-missing values. p-values are from Pearson’s chi-squared test or Fisher’s exact test, as appropriate. ^1^n (%). ^2^Pearson’s chi-squared test; Fisher’s exact test. ^3^Histology: non-squamous, squamous, or other. The "other" category comprises undifferentiated carcinoma, small-cell lung carcinoma, neuro-endocrine carcinoma, and unknown or missing entries. ^4^Mutation frequencies are reported among patients with available whole-exome sequencing data. Additional mutations identified but not listed in the table include: HER2 (16), CTNNB1 (17), DDR2 (14), FBXW7 (13), SMAD4 (17), ERBB4 (27), BRAF (31), FGFR2 (15), FGFR3 (15), PDGFRA (26), PTEN (27), MET (18), TERT (13), NRAS (7), PIK3CA (28), EGFR (13), KIT (21), POLE (26), RET (22), HRAS (1), IDH1 (1), KIF5B (1), ERBB2 (2), ROS1 (19), TTF1 (1), ANTI_CYTOKERATINE (1), CDKN2A (24). CT, chemotherapy; ICI, immune checkpoint inhibitor; ECOG, Eastern Cooperative Oncology Group; PD, progressive disease; SD, stable disease; PR, partial response; CR, complete response; N/A, not evaluable; PD-L1, programmed death-ligand 1; TPS, tumour proportion score.

| Table 1: Patient characteristics |  |  |  |  |
| --- | --- | --- | --- | --- |
| Variable | Overall, N = 439<sup>1</sup> | Treatment setting |  | p-value<sup>2</sup> |
|  |  | 1L CT+ICI, N = 269 | 2L+ ICI, N = 170 |  |
| Age category |  |  |  | <0.001 |
| <70 | 323 (74%) | 214 (80%) | 109 (64%) |  |
| ≥70 | 115 (26%) | 54 (20%) | 61 (36%) |  |
| Missing | 1 | 1 | 0 |  |
| Histology<sup>3</sup> |  |  |  | 0.003 |
| Non-squamous | 314 (72%) | 198 (74%) | 116 (68%) |  |
| Other | 52 (12%) | 38 (14%) | 14 (8.2%) |  |
| Squamous | 73 (17%) | 33 (12%) | 40 (24%) |  |
| Liver metastases |  |  |  | 0.041 |
| No | 373 (85%) | 236 (88%) | 137 (81%) |  |
| Yes | 66 (15%) | 33 (12%) | 33 (19%) |  |
| ECOG |  |  |  | <0.001 |
| 0 | 177 (40%) | 126 (47%) | 51 (30%) |  |
| 1 | 262 (60%) | 143 (53%) | 119 (70%) |  |
| Sex |  |  |  | >0.9 |
| Female | 153 (35%) | 94 (35%) | 59 (35%) |  |
| Male | 286 (65%) | 175 (65%) | 111 (65%) |  |
| Type of drug |  |  |  | <0.001 |
| Atezolizumab | 53 (12%) | 0 (0%) | 53 (31%) |  |
| Nivolumab | 51 (12%) | 0 (0%) | 51 (30%) |  |
| Pembrolizumab | 66 (15%) | 0 (0%) | 66 (39%) |  |
| Pembrolizumab + chemotherapy | 269 (61%) | 269 (100%) | 0 (0%) |  |
| Smoking history |  |  |  | 0.2 |
| Current | 154 (36%) | 100 (38%) | 54 (33%) |  |
| Never | 24 (5.6%) | 11 (4.2%) | 13 (7.8%) |  |
| Previous | 250 (58%) | 151 (58%) | 99 (60%) |  |
| Missing | 11 | 7 | 4 |  |
| Primary resistance |  |  |  | <0.001 |
| No | 263 (60%) | 188 (70%) | 75 (45%) |  |
| Yes | 172 (40%) | 80 (30%) | 92 (55%) |  |
| Missing | 4 | 1 | 3 |  |
| Best Overall Response |  |  |  | <0.001 |
| PD | 45 (10%) | 9 (3.3%) | 36 (21%) |  |
| SD | 141 (32%) | 76 (28%) | 65 (38%) |  |
| PR | 154 (35%) | 128 (48%) | 26 (15%) |  |
| CR | 23 (5.2%) | 15 (5.6%) | 8 (4.7%) |  |
| N/A | 76 (17%) | 41 (15%) | 35 (21%) |  |
| Mutations<sup>4</sup> |  |  |  |  |
| KRAS | 130 (42%) | 87 (42%) | 43 (41%) |  |
| TP53 | 173 (56%) | 117 (57%) | 56 (54%) |  |
| STK11 | 58 (19%) | 40 (20%) | 18 (17%) |  |
| KEAP1 | 53 (17%) | 41 (20%) | 12 (12%) |  |
| PD-L1 expression |  |  |  | 0.010 |
| <1% | 200 (55%) | 110 (50%) | 90 (63%) |  |
| [1%, 50%] | 77 (21%) | 45 (21%) | 32 (22%) |  |
| >50% | 85 (23%) | 63 (29%) | 22 (15%) |  |
| Missing | 77 | 51 | 26 |  |

<sup>1</sup> n (%)
<sup>2</sup> Pearson's Chi-squared test; Fisher's exact test
<sup>3</sup> Histology. Can either be 'Non-squamous', 'Squamous' or 'Other'. Category 'Other' consists of original entries 'UNK: Unknown', 'UC: Undifferentiated carcinoma', 'SCLC: small cell lung carcinoma', 'NEC: neuro-endocrine carcinoma', 'OTHER', 'NSCLC' and missing values.
<sup>4</sup> Other hidden mutations are: HER2 (16), CTNNB1 (17), DDR2 (14), FBXW7 (13), SMAD4 (17), ERBB4 (27), BRAF (31), FGFR2 (15), FGFR3 (15), PDGFRA (26), PTEN (27), MET (18), TERT (13), NRAS (7), PIK3CA (28), EGFR (13), KIT (21), POLE (26), RET (22), HRAS (1), IDH1 (1), KIF5B (1), ERBB2 (2), ROS1 (19), TTF1 (1), ANTI\_CYTOKERATINE (1), CDKN2A (24)

### Multimodal pretreatment profiling captured 442 features across six biological layers

Comprehensive multimodal profiling of the pretreatment baseline timepoint yielded 442 individual biomarker features spanning six biological layers — tumour microenvironment, tumour molecular profiling, circulating immune cells, vasculo-monitoring, routine haematology and biochemistry, and standard clinical variables — of which 394 were continuous and 48 categorical (Figure 1B-C, Supplementary Table 1). The overall missing-value rate was 39% (Figure 1D), reflecting the practical challenges of deep multimodal phenotyping in real-world prospective settings. Missingness was strongly layer-dependent, which was investigated in another, methodological, work ^44^.

Correlation analyses confirmed the expected within-layer biological structure but limited correlation between circulating and tumour compartments, supporting the rationale for multimodal integration (interactive reports: https://compo.inria.fr/pioneer-website/technical-reports.html).

The analytical workflow began with filtering of high-missingness features and iterative VIF-based collinearity reduction, yielding a decorrelated dataset of 214 features. Within a .632 optimism-correction framework, 36 feature-selection methods were combined with 10 machine-learning models and benchmarked for predictive performance and stability, with downstream analyses of the resulting signature, PFS stratification, and individual-level SHAP-based interpretation (Figure 1D)

### No single biomarker strongly discriminated primary resistance

After PD-L1 adjustment and FDR correction, 14 biomarkers were significantly associated with PrR, 33 with PFS, and 39 with OS (q<0.05, decorrelated feature set), with 13 markers reaching significance across all three endpoints (Figure 2A, Table 2; Supplementary Figure 3). All PrR-associated markers were also associated with PFS or OS, with no PrR-specific signals.

**Figure 2:**
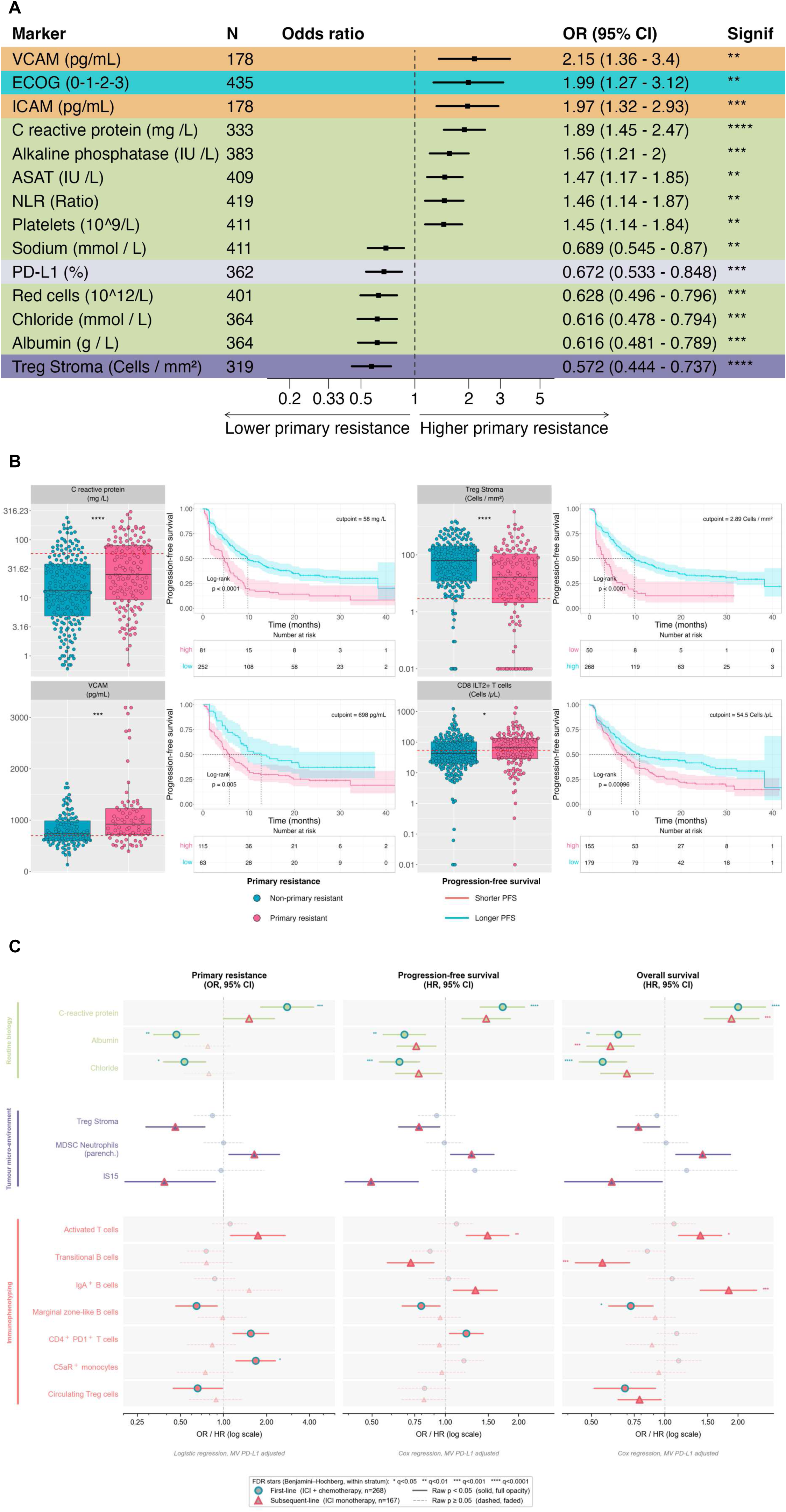
Single markers. A. Forest plot of markers significantly associated with primary resistance (PD-L1-controlled logistic regression, FDR-adjusted q-value < 0.05). Decorrelated dataset. The number of patients with a non-missing value is given for each marker. B. Box and Kaplan-Meier (KM) plots of the most significant features for PFS (log-rank test), for each type of biomarkers. Red dashed line = KM optimal cutoff. C. Treatment-line-stratified associations of selected biomarkers with primary resistance, progression-free survival, and overall survival. For each biomarker, results from first-line (ICI+CT, n=268; circles) and subsequent-line (ICI, n=167; triangles) patients are displayed as paired rows. Point fill colour indicates biological modality: green = routine biology; purple = tumour micro-environment; red = immunophenotyping. The edge colour of each symbol encodes treatment line: teal = first-line; salmon = subsequent-line. Horizontal bars represent 95% confidence intervals. Full opacity and solid bars indicate raw p < 0.05; faded symbols and dashed bars indicate p ≥ 0.05. Significance stars are derived from FDR-adjusted q-values (Benjamini–Hochberg correction applied independently within each treatment-line stratum): *q<0.05, **q<0.01, ***q<0.001, ****q<0.0001. All models are multivariable, adjusted for PD-L1 expression as a continuous covariate. Primary resistance was assessed by logistic regression (odds ratio, OR); progression-free survival and overall survival by Cox proportional-hazards regression (hazard ratio, HR). Markers are grouped by biological modality and ordered within each group from line-2+-dominant (upper rows) to line-1-dominant (lower rows) associations. MDSC: myeloid-derived suppressor cells; IS15: Isign 15-gene immune contexture transcriptomic signature; IgA: immunoglobulin A; C5aR: complement component 5a receptor; Treg: regulatory T cells. CI: confidence interval. Unless expressed differently, all statistics are from PD-L1-controlled logistic/Cox regressions, FDR-adjusted q-value < 0.05 on the decorrelated dataset. VCAM: Vascular Cell Adhesion Molecule; ICAM: Intercellular Adhesion Molecule. See the dashboard for direct exploration of all individual biomarkers at https://compo.inria.fr/pioneer-dashboard/.

**Table 2:**
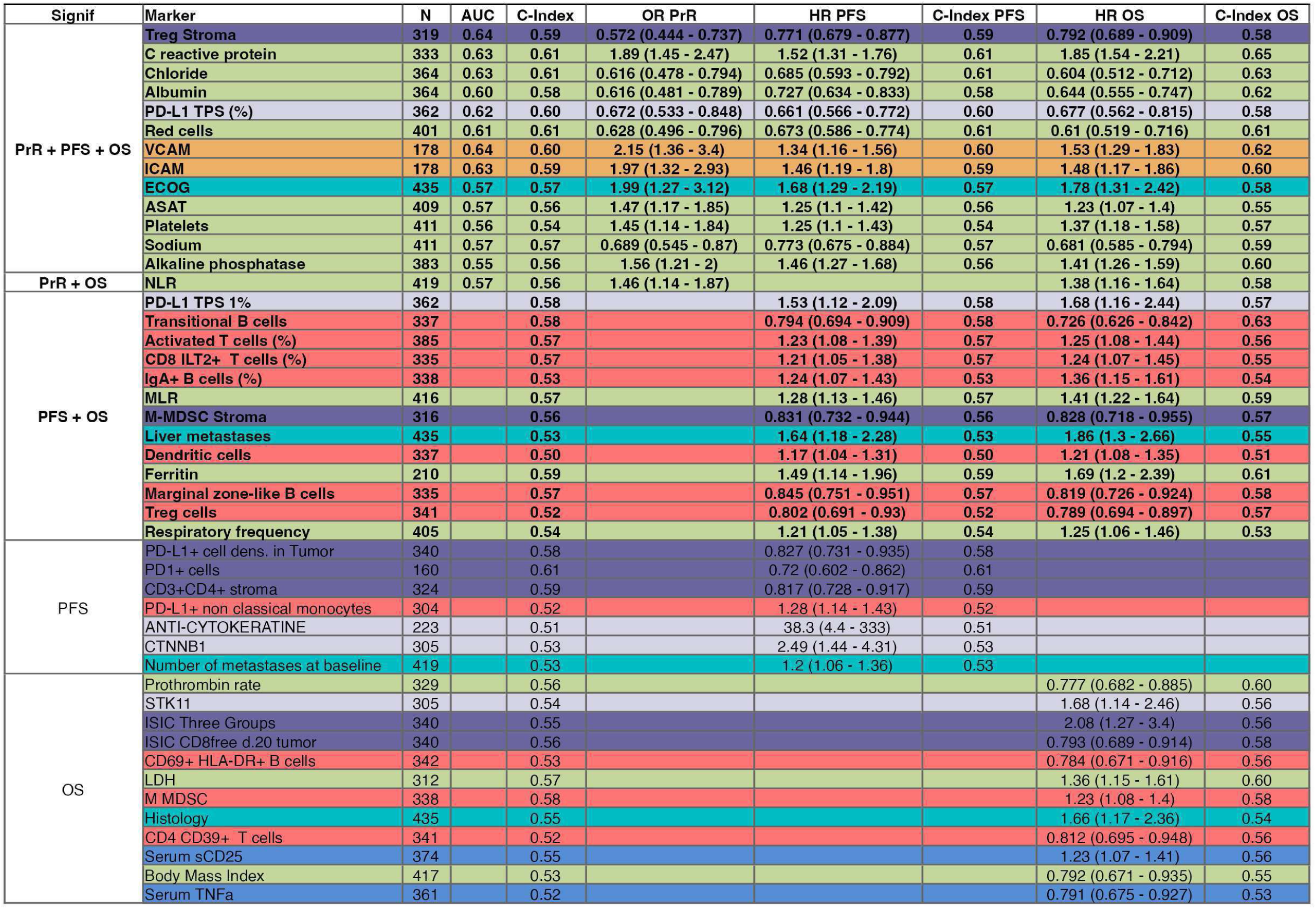
Single biomarkers statistics. Biomarkers reaching statistical significance (PD-L1-controlled, FDR-adjusted q ≤ 0.05) for at least one of three endpoints, grouped by the combination of endpoints they reach: primary resistance (PrR; multivariable logistic regression), progression-free survival (PFS; multivariable Cox regression), and overall survival (OS; multivariable Cox regression). AUC denotes the area under the ROC curve for PrR; C-Index denotes the concordance index from the Cox model for PFS; OR and HR denote the multivariable odds ratio and hazard ratios for PrR, PFS, and OS, respectively. Row background colors indicate biomarker category (see Figure 1). N denotes the number of patients with a non-missing value for that marker. PD-L1: pathologist-assessed TPS score.

No single biomarker showed strong discrimination for PrR (Figure 2A, Table 2). The highest individual AUCs were for stromal Treg infiltration, blood VCAM, and blood CRP (all ≤ 0.64), which also ranked among the strongest PFS predictors. PD-L1 TPS retained a significant association with both PrR (AUC 0.62) and PFS (C-index 0.60), but remained only modestly discriminatory; among patients with PD-L1 TPS below 1%, 52% nonetheless met criteria for primary resistance. TMB was non-discriminatory for both PrR (AUC 0.55) and PFS (C-index 0.54).

Notable findings included a discrete subgroup with no measurable stromal immune infiltration or with elevated VCAM (≥ 2,000 pg/mL), all of whom met PrR criteria (Figure 2B). Several other markers reached FDR significance, including stromal PD-1⁺ cell density, circulating CD8⁺ILT2⁺ Supplementary Figure 4). T cells, and circulating transitional B cells (Table 2, Among genomic alterations, no single mutation reached FDR significance for PrR. *STK11* mutation was associated with worse PFS in both treatment settings (p = 0.015 and 0.018, log-rank), *KEAP1* mutation in the first-line setting only (p = 0.018), and *CTNNB1* mutations were strongly enriched among very-early progressors despite low overall frequency.

Analyses of secondary binary endpoints identified 10 markers associated with objective response — dominated by circulating NK-cell and T-cell activation features — and 25 with early progression, the richest secondary endpoint in terms of significant associations (Supplementary Figure 5).

Complete results for all biomarkers are available on the study dashboard (https://compo.inria.fr/pioneer-dashboard/).

### Treatment-setting stratification revealed divergent association patterns

In first-line patients receiving chemo-immunotherapy (n=268), PrR and PFS associations were dominated by routine blood chemistry and systemic inflammatory markers (Figure 2C, Supplementary Table 2). Eight markers survived FDR correction for PrR, all from the routine biology domain, alongside circulating C5aR⁺ CD14*Low* CD16⁻ monocytes — the only circulating immune marker reaching FDR significance in this setting. Tumour microenvironment markers — including stromal Treg cell density — showed no meaningful association with any endpoint in first-line patients.

In subsequent-line patients receiving ICI monotherapy (n=167), the pattern was reversed: no routine blood marker survived FDR correction, and dominant associations shifted to immunophenotyping and tumour microenvironment variables, with consistent directional effects across PrR, PFS, and OS for stromal Treg infiltration, parenchymal neutrophils, ImmunoSign 15, circulating activated T cells, transitional B cells, and IgA⁺ B cells (Supplementary Table 2). This biological dissociation between treatment settings likely reflects the confounding influence of concurrent chemotherapy on immune baseline measurements in the combination setting.

Single-biomarker analyses thus revealed a broad but individually modest association landscape — no marker exceeded AUC 0.64 for PrR — and highlighted the orthogonality of circulating and tumour compartment signals, motivating multimodal integration.

### Multimodal machine learning integration outperformed PD-L1 and TMB and stratified PFS

To maximise statistical power while preserving treatment-setting heterogeneity, data from first-line and subsequent-line patients were pooled (n=435), with treatment line retained as a predictor. Thirty-six feature-selection methods were combined with ten machine-learning models, yielding 360 pipeline combinations (Figure 1D). Across all methods, stability and optimism-corrected AUC were correlated, with BH-filtered PFS-Cox regression (q < 0.01) occupying the Pareto frontier of both dimensions (Supplementary Figure 6); this method was carried forward as the minimal multimodal model^34,39^.

Applied to the full 214-feature decorrelated dataset, the multimodal pipeline achieved an optimism-corrected AUROC of 0.73 ± 0.05, PPV of 0.63 ± 0.07, and F1 of 0.60 ± 0.06, with adequate calibration on out-of-bag predictions (Supplementary Figure 7A-B). The pipeline incorporating the BH-filtered PFS-Cox feature selection step showed comparable discrimination with a more favourable sensitivity-specificity trade-off.

In treatment-setting-specific analyses, the multimodal model outperformed both PD-L1 TPS and TMB in the first-line CT+ICI setting, with a more balanced metric profile (PPV 0.51 ± 0.11 versus 0.36 ± 0.02 and 0.35 ± 0.03; Figure 3A). Decision curve analysis confirmed clinical utility: the multimodal model yielded higher net benefit than both PD-L1 and treat-all/treat-none strategies across a wide range of decision thresholds (Figure 3B). In the subsequent-line monotherapy setting, the multimodal model did not outperform PD-L1. Although its AUROC was marginally higher (0.69 ± 0.08 versus 0.66), PD-L1 alone was numerically superior on the study’s co-primary metric PPV (0.68 versus 0.64) and, unlike the model, a single pre-specified marker involves no fitting and so carries no optimism at all. A plausible interpretation is that, in the chemotherapy-free setting, tumour PD-L1 retains more of its predictive content, whereas in the combination setting part of the outcome is attributable to chemotherapy and dilutes it. The smaller sample (n=167) also limits what can be resolved.

**Figure 3:**
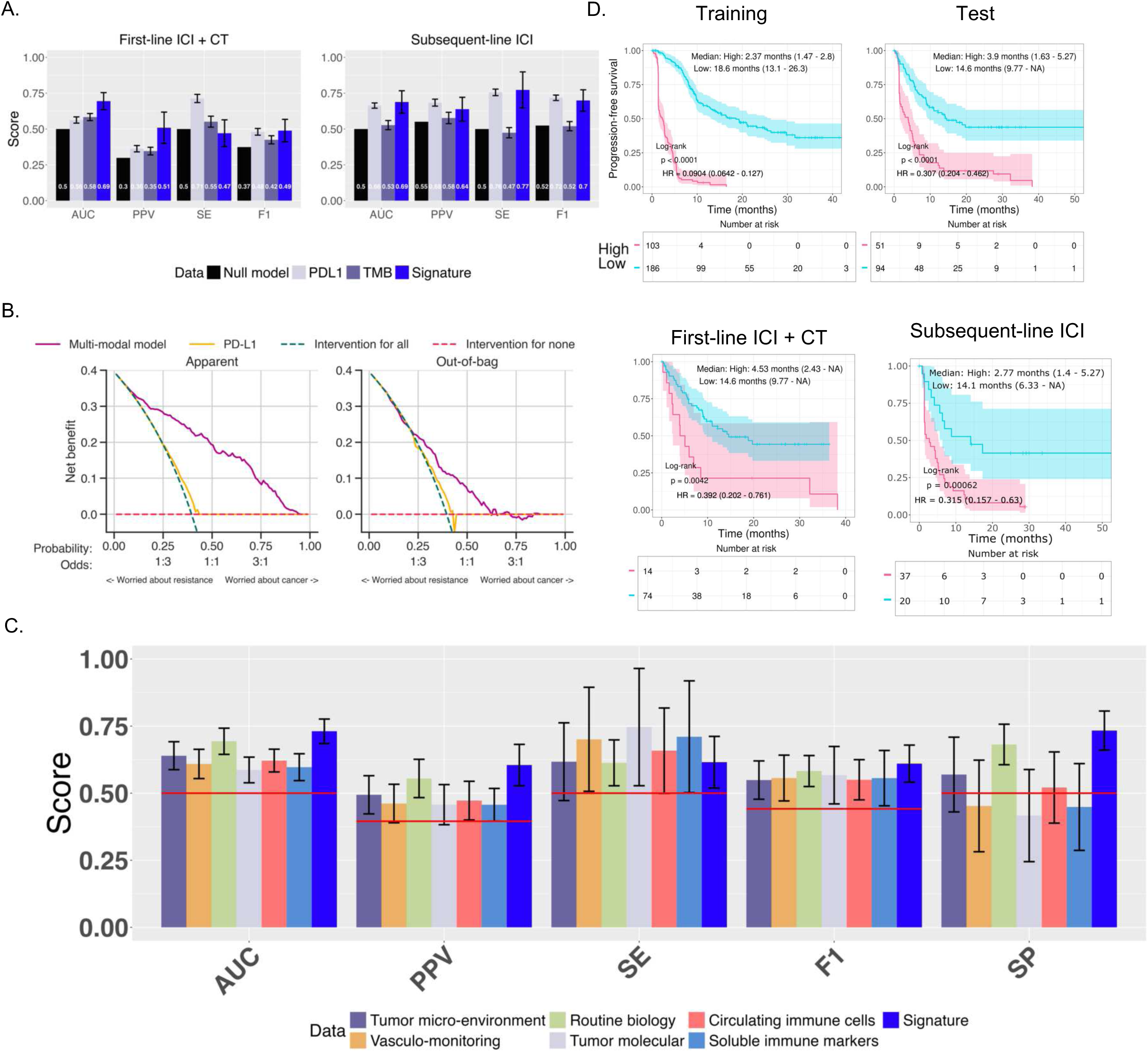
Multimodal machine learning predictive performance. A. Treatment-setting-stratified optimism-corrected performance metrics for the multimodal model, shown separately for first-line ICI+CT (left) and subsequent-line ICI (right). Bar colours represent the null model (black), PD-L1 (light grey, 1% threshold for all metrics except AUC where the continuous marker was used), TMB (dark grey, 10 mutations/megabases threshold for all metrics except AUC where the continuous marker was used), and the multimodal signature (dark blue). B. Decision curve analysis for the multimodal model and PD-L1 across a range of treatment thresholds. Net benefit is plotted as a function of probability threshold (top axis) and corresponding odds (middle axis) for the apparent (left) and out-of-bag (right) estimates. The multimodal model (solid magenta line) and PD-L1 expression alone (solid orange line) are compared against the two reference strategies: treat all patients (teal dashed line) and treat no patients (red dashed line, net benefit = 0). The lower x-axis indicates the clinical interpretation of the threshold: low thresholds correspond to scenarios where the clinician is primarily concerned about missing resistance (left), while high thresholds reflect greater concern about undertreating cancer (right). A model provides clinical utility when its net benefit curve lies above both reference lines. The out-of-bag panel reflects cross-validated performance and therefore represents a less optimistic, more generalizable estimate of clinical utility. C. Comparison of optimism-corrected predictive metrics across uni-modal models and the multimodal model. Each bar represents performance from a pipeline trained exclusively on the indicated data modality. The red line denotes the null model baseline. D. Progression-free survival stratification by the multimodal risk score, shown for the training set (top left) and independent test set (top right), and stratified by treatment line: first-line ICI+CT (bottom left) and subsequent-line ICI (bottom right). Patients were classified as high-risk (pink) or low-risk (teal) based on a cutpoint derived in the training set and propagated to the test set. Median PFS and hazard ratios with 95% confidence intervals are shown; p-values are from the log-rank test. The cutpoint was derived in the training set, so the training-set hazard ratio is an apparent value; the test-set hazard ratio is the internally validated one. AUC, area under the receiver operating characteristic curve; PPV, positive predictive value; SE, sensitivity; SP, specificity; F1, F1 score; ICI, immune checkpoint inhibitor; CT, chemotherapy; PFS, progression-free survival; HR, hazard ratio; CI, confidence interval; TMB, tumour mutational burden; PD-L1, programmed death-ligand 1; BH, Benjamini–Hochberg.

When benchmarked against grouped biomarker modalities, the multimodal model outperformed unimodal models across several performance metrics, particularly PPV (Figure 3C). Among unimodal data blocks, routine laboratory blood tests were the most informative, highlighting the underused predictive value of clinically accessible baseline variables.

Although trained on binary PrR, the model also stratified PFS (Figure 3D). In the training cohort, median PFS was 2.37 months (95% CI 1.47–2.8) in high-risk patients versus 18.6 months (13.1–26.3) in low-risk patients (apparent Hazard Ratio (HR) 0.090, 95% CI 0.064–0.127; log-rank p<0.0001). Similar separation was observed in the independent test set (3.9 versus 14.6 months; HR 0.31, 95% CI 0.20–0.46; p<0.0001) and was maintained across both treatment settings.

### An 18-feature signature revealed heterogeneous biological routes to primary resistance

Feature selection on the decorrelated dataset yielded an 18-biological-variable signature plus treatment line, spanning pathological, immunological, vascular, and routine laboratory layers (Figure 4A). Among the most influential features by SHAP global importance were stromal Treg infiltration, PD-L1 expression, and circulating transitional B cells, alongside multiple routine laboratory variables. Routine blood parameters collectively accounted for approximately half of the final signature, with blood chloride showing the strongest SHAP global importance, followed by albumin, red blood cell count, monocyte-to-lymphocyte ratio, alkaline phosphatase, and CRP. Chloride also defined the first split of the model-derived decision tree (Figure 4B), in which treatment setting, circulating transitional B cells, stromal Treg infiltration and PD-L1 expression formed the subsequent branches.

**Figure 4.**
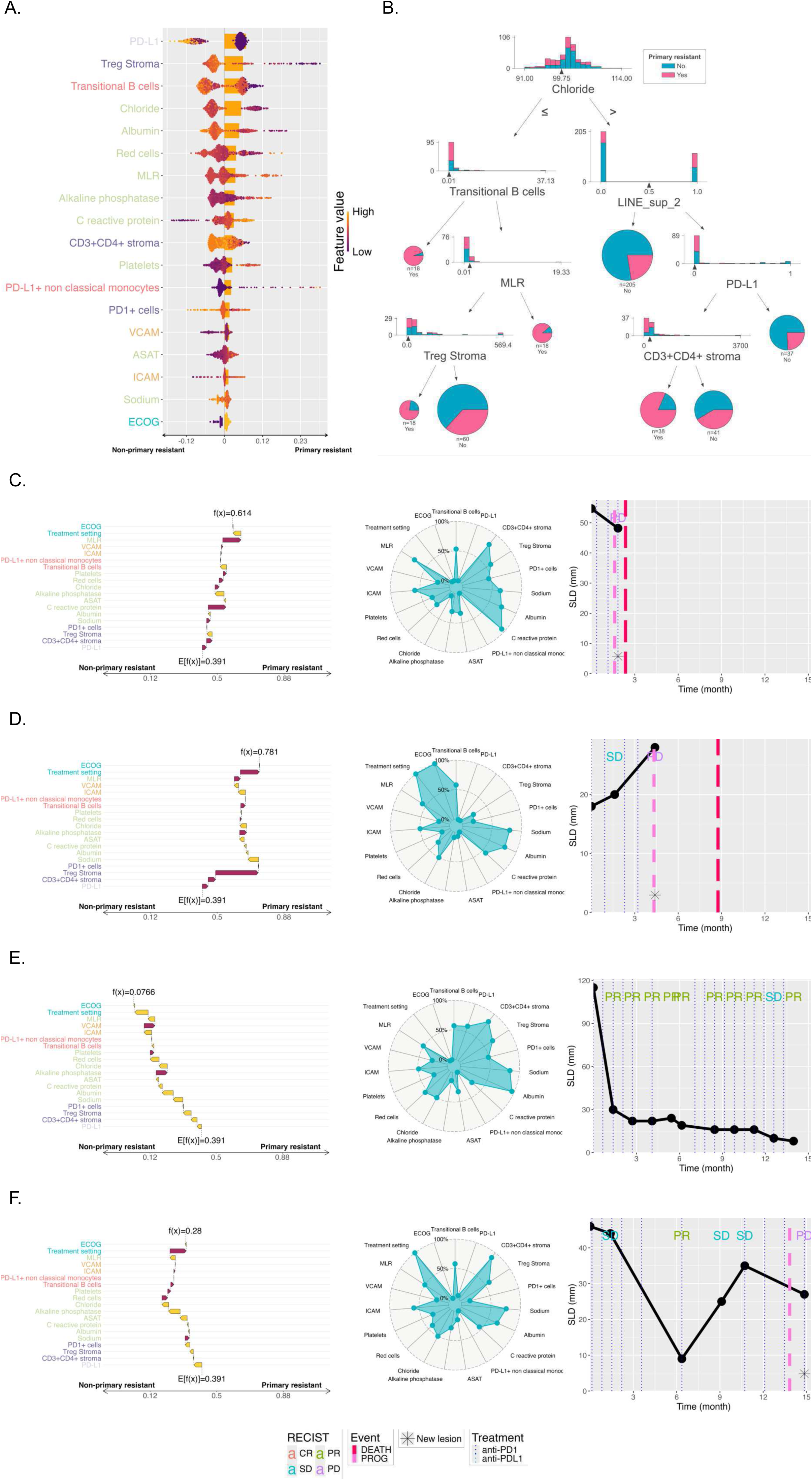
Multimodal signature, decision tree, and patient-level interpretation. A. Multimodal signature derived from feature selection on the decorrelated biomarker dataset, sorted by gradient boosting SHAP global importance. Each point represents the SHAP value of one patient for one signature variable, coloured by the patient’s value of that variable from low (yellow) to high (purple). Treatment line was retained as a covariate but is not displayed. B. Decision trees fitted on signature variables. Histograms show the distribution of each split variable in primary-resistant (pink) and non-primary-resistant (teal) patients, with triangles indicating the cutpoints; pie charts at terminal nodes show the proportion of primary-resistant patients reaching that node, with node sizes proportional to patient count. For Treg Stroma, the histogram was truncated at the 95th percentile for visualisation. C–F. Patient-level interpretation of the multimodal signature for four representative correctly predicted patients in the test set. Left: SHAP waterfall plot showing the contribution of each signature variable to the predicted probability of primary resistance, starting from the population baseline E[f(x)] = 0.391 and ending at the individual prediction f(x). Middle: radar plot of the patient’s signature variable values, expressed as percentiles within the overall cohort. Right: time course of the sum of longest diameters with RECIST 1.1 assessments and progression or death events. C. A first-line ICI+CT primary-resistant patient (f(x) = 0.614) progressing within 3 months despite immune infiltration; resistance was driven primarily by a systemic inflammatory profile (elevated CRP and MLR). D. A subsequent-line ICI primary-resistant patient (f(x) = 0.781) with progression after stable disease at ∼5 months; resistance was driven primarily by low stromal Treg, concomitant with non-contributive but low PD-1⁺ cells and low CD3⁺CD4⁺ infiltration. E. A first-line ICI+CT non-primary-resistant patient (f(x) = 0.077) with sustained partial responses and one complete response across more than a year of follow-up; favourable classification was driven by contributions from almost all markers of the signature, with a notably low contribution of high PD-L1 TPS. F. A subsequent-line ICI non-primary-resistant patient (f(x) = 0.28) with durable stable disease and a partial response over 13 months despite low PD-L1 expression; protective favourable medical biology. RECIST: Response Evaluation Criteria In Solid Tumours; SLD: sum of longest diameters; PD: progressive disease; SD: stable disease; PR: partial response; CR: complete response; SHAP: SHapley Additive exPlanations; MLR: monocyte-to-lymphocyte ratio; VCAM: vascular cell adhesion molecule; ICAM: intercellular adhesion molecule; ASAT: aspartate aminotransferase; CRP: C-reactive protein; ECOG: Eastern Cooperative Oncology Group performance status; TPS: tumour proportion score.

Hierarchical clustering of patients on signature variables revealed a coherent pattern in which primary-resistant patients (PrR = 1) showed elevated systemic inflammation markers (platelets, CRP, alkaline phosphatase, aspartate aminotransferase (ASAT)) together with depressed nutritional and homeostatic parameters (albumin, sodium, chloride, red cells), consistently across both first-line and subsequent-line settings (Supplementary Figure 9). In the subsequent-line setting, a subset of primary resistant patients additionally exhibited markedly reduced stromal Treg and CD3+CD4+ T-cell infiltration and low circulating transitional B-cell counts, recapitulating the survival associations identified in univariable analyses. Blood VCAM and ICAM showed moderate elevation in primary resistant patients, more visibly in the monotherapy setting. By contrast, tumor PD-L1 expression showed no cluster-level enrichment, reflecting its heterogeneous and individually modest contribution to resistance classification.

Patient-level SHAP decomposition in the test set revealed marked inter-patient heterogeneity in the relative contributions of these features (Figure 4C-F). In some PrR patients, CRP (Figure 4C) or low stromal Treg infiltration (Figure 4D) were the primary risk drivers, preceding low PD-L1 TPS. Among durable responders, some were classified as low-risk primarily through high PD-L1 TPS, whereas others with low PD-L1 expression had favourable prediction resulting from multiple contributive markers (Figure 4E-F). These profiles support the biological heterogeneity of primary resistance and reinforce the case for multimodal rather than single-biomarker risk assessment.

## DISCUSSION

In this prospective multicentre biomarker study, comprehensive multimodal profiling revealed a broad but individually modest landscape of associations with primary resistance to PD-(L)1 inhibitors, progression-free survival, and overall survival in advanced NSCLC patients. Multimodal integration of routine and specialised biomarkers achieved a meaningful predictive signal, with an optimism-corrected PPV of 51% in the first-line ICI+CT setting, without easily derivable clinical consequences for daily practice however. Nonetheless, the developed signature could be used to identify advanced NSCLC patients appropriate for next generation immunotherapy clinical trials. Furthermore, complete results for all 442 profiled biomarkers are publicly accessible through a companion website, in support of open science and positioning PIONeeR as a reference dataset for future immune-oncology biomarker meta-analyses and external validation efforts.

Significant biomarkers were dominated by routine blood-based biology, suggesting that a substantial fraction of the resistance signal is accessible from usual pretreatment tests and potentially reflecting a greater value of the systemic, compared with the tumoural or microenvironmental, component of the immune system in explaining and predicting resistance. As an illustration, among routine biology variables, the prominence of chloride likely reflects a broader homeostatic axis rather than a chloride-specific effect, with hypochloremia covarying with hyponatremia and hypoalbuminemia in the primary-resistant cluster (Supplementary Figure 9), consistent with cancer-associated cachexia and systemic inflammation. Chloride was independently identified as a top prognostic feature in the large-scale SCORPIO model^25^, supporting its relevance as a robust, routinely available marker of immunotherapy outcome. Within the tumour microenvironment, stromal Treg assessment showed a counterintuitive association with primary resistance: the complete absence of measurable stromal Treg infiltration, rather than high Treg counts, more strongly predicted the risk of primary resistance. While tumour-infiltrating Tregs are canonically associated with immunosuppression and poor outcomes in NSCLC^45^, this finding can be interpreted through the immune-desert framework^46,47^. Among circulating immune populations, CD8⁺ILT2⁺ T cells expressing an inhibitory receptor that suppresses TCR-mediated cytotoxicity were associated with shorter PFS and OS; ILT2 (LILRB2) is an emerging therapeutic target currently in early-phase trials, lending translational relevance to this observation^48^. Elevated blood soluble VCAM-1 was associated with primary resistance across endpoints, plausibly reflecting endothelial dysfunction and impaired immune-cell trafficking to the tumour^49^. The biological dissociation between treatment settings — routine inflammatory and metabolic markers accounting for most significant associations in first-line patients, tumour immune contexture and circulating lymphocyte subsets accounting for most in subsequent-line patients — suggests that biomarker relevance is not setting-agnostic. Altogether, these findings provide new hypotheses for strategies aimed at overcoming resistance to PD-(L)1 blockade.

In the first-line ICI+CT setting, 30% of patients met primary-resistance criteria, so flagging patients at random would yield a PPV of 30%. The multimodal model’s optimism-corrected PPV of 51% therefore represents a meaningful improvement over this prevalence-determined baseline, and over the 36% achieved by PD-L1 TPS at the 1% threshold. Unlike AUC or C-index, PPV has a straightforward clinical interpretation: more than half of model-predicted PrR patients would indeed progress within 6 months. The substantial contribution of routine laboratory variables to this performance is consistent with recent findings^25^.

While several previous studies performed multimodal integration — ranging from deep multi-platform phenotyping^19,22,23,50^ to shallow-feature large-cohort models built on routine clinical and laboratory data^24,25^ — PIONeeR is the only study to have systematically profiled six biological layers — clinical, routine laboratory, circulating immune cells, soluble vascular markers, tumour immune contexture by multiplex IHC and IF, and genomics — in a prospective, multicentre cohort spanning both treatment settings (Supplementary Table 3). Direct comparison is hampered by heterogeneity across endpoints, populations, and methodology. Most studies use ORR^19,23,24^, which is not equivalent to SITC-defined primary resistance^26,27^; the two studies reporting durable clinical benefit^22,25^ exhibited lower AUROC (0.67 ± 0.03^22^ and 0.63^25^ versus 0.73 ± 0.05 in PIONeeR), with no report of more clinically meaningful metrics such as PPV or net benefit curves. Most models were developed exclusively in monotherapy-treated patients^19,23,24^; fully multimodal studies had limited effective sample sizes (n = 81^23^, 80^22^). Methodological standardisation is largely absent: feature-selection stability across subsamples is rarely evaluated^51^, and adherence to TRIPOD recommendations^52^ — which mandate optimism-corrected internal validation — remains uncommon. PIONeeR addressed these gaps by benchmarking 36 feature selection methods, embedding the full modelling pipeline within each resample, and applying .632 optimism correction throughout to control overfitting^37,38^.

Some limitations should however be acknowledged. First, PIONeeR lacks an independent external validation cohort; optimism-corrected internal validation provides a principled estimate of generalisation error but does not substitute for testing in a population with a potentially different distribution of patient and biomarker characteristics. The performance reported here therefore carries no easily derivable consequence for daily practice yet, and should be read as a benchmark for what deep multimodal pretreatment profiling can achieve rather than as a deployable decision rule. Second, all analyses were restricted to static pretreatment measurements, although paired on-treatment samples were collected and will be the subject of future analyses. Third, several biomarker layers — flow cytometry-based immune phenotyping, multiplex IHC and IF, and whole-exome sequencing — are not routinely available across centres, limiting immediate clinical translatability of the full signature. Fourth, the absence of a non-ICI comparator arm means that predictive and prognostic effects cannot be fully disentangled. Finally, the predictive signal reported here is derived from a defined set of biological layers, chosen for their direct interpretability. Several additional modalities profiled in PIONeeR – including full transcriptomics, circulating tumour nucleic acids, and gut microbiome composition – were not integrated into the present analyses and are reserved for future work, while others not assessed in this study, such as radiomics, pathomics, and spatial transcriptomics, may carry complementary information on resistance. Their integration could further refine pretreatment risk prediction. Three methodological caveats should finally be weighed. The reported pipeline was selected from 360 candidates on optimism-corrected performance, so its metrics carry a winner’s-curse bias; the distribution across the whole grid is given in Supplementary Figure 6. The overall missing-value rate was 39% and varied by an order of magnitude between biological layers, so median-imputed values contributed materially to the model for the least-complete markers. And with 172 events for 214 candidate predictors, the discovery step is data-poor by prediction-modelling standards, even though the 19-variable final model corresponds to a more conventional 9.1 events per variable.

Taken together, these findings establish that primary resistance to anti–PD-(L)1 therapy in advanced NSCLC is a composite biological state that cannot, as expected, be captured by any single marker, but is meaningfully predicted by multimodal integration of pretreatment variables. The important contribution of routine blood-based parameters to the predictive signal is of immediate practical relevance: it suggests that a clinically implementable simplified score — prospectively validated and benchmarked against existing risk stratifications — could identify patients unlikely to benefit from immunotherapy before treatment initiation, informing more individualised therapeutic decision-making.

In addition, the identified biomarkers provide hypotheses regarding potential targets for future strategies aimed at overcoming primary resistance to PD-(L)1 inhibition in advanced NSCLC.

## Data Availability

All data produced in the present study are available upon reasonable request to the authors

## DATA AVAILABILITY

Aggregate results for all 442 profiled biomarkers — univariable distributions, associations with every endpoint, and the full statistical and machine-learning reports — are publicly available without restriction through the study dashboard (https://compo.inria.fr/pioneer-dashboard/) and the companion technical-report site (https://compo.inria.fr/pioneer-website/). Individual patient-level clinical and biomarker data are not publicly available because participants did not consent to unrestricted public release and the data remain subject to the French and EU data-protection framework (GDPR) under the sponsorship of Assistance Publique – Hôpitaux de Marseille. De-identified individual-level data may be made available to qualified investigators for a pre-specified research purpose, following approval by the PIONeeR scientific committee and execution of a data-transfer agreement with the sponsor. Requests should be addressed to.

## CODE AVAILABILITY

The three analysis packages developed for this study are open source: compo.EDA (https://gitlab.inria.fr/compo/compo.eda), compo.tidyML (https://gitlab.inria.fr/compo/compo.tidyml) and ROOFS (RObust biOmarker Feature Selection, https://gitlab.inria.fr/compo/roofs) ^34^.

## ACKNOWLEDGEMENTS

This work benefited from a government grant handled by the French National Research Agency (ANR) as part of the France 2030 investment plan, under the reference ANR-17-RHUS-0007.

This work was supported by a partnership of Aix-Marseille Université (AMU), Assistance Publique Hôpitaux de Marseille (APHM), Centre National de La Recherche Scientifique (CNRS), Institut National de la Santé et de la Recherche Médicale (INSERM), Centre Léon Bérard (CLB), Institut Paoli Calmettes (IPC), Gustave Roussy (GR), AstraZeneca (AZ), Veracyte (VERA), Innate Pharma (IPH) & ImCheck Therapeutics (ICT), and initiated by Marseille Immunopole. The authors gratefully acknowledge the support of the APHM, which sponsored the PIONeeR clinical studies. Its role was to control the appropriateness of ethical and legal considerations for all centers and to perform the monitoring of the consents signed and the clinical data recorded and coded as part of the study.

The authors are grateful to all the patients and their families, as well as all the investigators, for their participation in the study.

This work benefited from support from ITMO Cancer AVIESAN and French Institut National du Cancer (grant #19CM148-00) and from the French National Research Agency (ANR), under the France 2030 program, reference ANR-22-PESN-0017.

## Supplementary Methods

### 1. Study design — detailed eligibility criteria and follow-up

Key eligibility criteria for the PIONeeR study included histologically confirmed advanced or recurrent NSCLC planned for treatment with standard-of-care anti–PD-(L)1 ICI. Patients receiving first-line therapy were scheduled for platinum-based chemotherapy plus anti–PD-(L)1 therapy, whereas patients receiving second- or third-line therapy had progressed on prior platinum-based chemotherapy and were scheduled for an approved anti–PD-(L)1 monotherapy. All patients had to have available archival formalin-fixed paraffin-embedded tumour tissue, be aged 18 years or older, have an Eastern Cooperative Oncology Group performance status of 0 or 1, have adequate organ function, and be anti–PD-(L)1-naive at study entry. Investigator-assessed radiographic tumour assessment was performed every 6 weeks using RECIST version 1.1^1^. Upon disease progression on ICI, patients entered a 24-week follow-up period during which subsequent anticancer therapy and survival data were collected (Supplementary Figure 1).

### 2. Endpoints — detailed evaluability rules

For monotherapy, primary resistance (PrR) was defined as investigator-assessed, RECIST 1.1^1^-based best overall response (BOR) of progressive disease or stable disease lasting less than 6 months from the first dose of ICI. For combination therapy, PrR was defined as progression within 6 months from the first dose of ICI irrespective of best overall response. To be evaluable for the primary endpoint, patients had to receive at least 6 weeks of treatment, except those with earlier progression in the combination-therapy setting. Secondary endpoints were progression-free survival (PFS), defined as time from study inclusion to disease progression or death; overall survival (OS), defined as time from study inclusion to death; objective response, defined as BOR being complete response or partial response versus stable or progressive disease; early progression, defined as progression within 3 months from the first dose of ICI; and long response, defined as no progression within 12 months from the first dose of ICI.

### 3. Clinical data and vital signs — detailed variables

Standard clinical data were collected within two weeks before ICI initiation and comprised 10 variables. These included patient characteristics (age, sex, smoking history), tumour-related features (histology, biopsy site, presence and number of metastatic sites including liver metastases, baseline sum of longest diameters), functional status assessed by the Eastern Cooperative Oncology Group performance status score, and baseline vital signs (heart rate, respiratory rate, and blood pressure). Treatment line — a key stratifying variable reflecting the distinct clinical contexts of first-line chemo-immunotherapy and subsequent-line monotherapy — was also recorded and retained as a covariate throughout all analyses. All variables were collected by the treating investigator as part of routine clinical assessment, requiring no additional patient procedures. The full list of variables is provided in Supplementary Table 1.

### 4. Routine haematology and biochemistry — detailed panel

Routine laboratory data were collected from standard-of-care blood sampling performed within two weeks before ICI initiation, with no additional patient procedures required for this purpose. The panel comprised 49 variables spanning a complete blood count with differential (including red blood cell count, haemoglobin, platelets, and absolute counts and derived ratios of neutrophils, lymphocytes, and monocytes), and a comprehensive serum biochemistry panel (including markers of hepatic function, renal function, electrolytes, inflammatory markers, and nutritional status). Derived inflammatory indices — including the neutrophil-to-lymphocyte ratio and monocyte-to-lymphocyte ratio — were computed from the differential count. All values were obtained from accredited hospital laboratories at each participating centre using locally validated assays, reflecting the standardised but real-world nature of these measurements. The full list of variables is provided in Supplementary Table 1.

### 5. Circulating immune profiling — full panel and timepoints

Peripheral blood samples were collected at baseline, prior to ICI initiation, for immune-cell phenotyping and soluble factor quantification. Immune-cell profiling was performed by flow cytometry on both fresh whole blood and thawed peripheral blood mononuclear cells, enabling characterisation of a broad range of circulating immune populations including T-cell, B-cell and NK cells subsets, monocytes, neutrophils, dendritic cells, and myeloid-derived suppressor cells, along with activation, exhaustion, and checkpoint expression states. Soluble immune mediators were quantified from serum using two complementary platforms: soluble CD25 was measured by ELISA, and a panel of key cytokines — interferon-gamma, tumour necrosis factor, IL-6, and IL-10 — was quantified using cytometric bead arrays. In total, this layer contributed 156 blood-based features to the multimodal dataset.

Samples collected on treatment were also available but were not used in the primary predictive analyses, which were restricted to the pretreatment baseline timepoint. The full panel of immune populations and soluble mediators profiled is detailed in Supplementary Table 1.

### 6. Multiplex immunohistochemistry — platforms, detection systems, and quantification

Archival tumour specimens were processed into up to 22 consecutive 4-micron FFPE sections per block. Immunohistochemistry (IHC) was performed according to a prespecified prioritisation scheme. Samples containing fewer than 100 viable tumour cells on the first section were considered non-evaluable. PD-L1 expression was assessed using the Immunoscore IC kit clone HDX3 (Veracyte) on Ventana Benchmark XT (Roche) or Bond RX (Leica) platforms, and tumour proportion score (TPS) was calculated as the percentage of viable tumour cells with membranous staining. Multiplex IHC panels targeting T-cell (CD3, CD8, PD-1, LAG-3, TIM-3) and myeloid-cell (CD11b, CD14, CD15, HLA-DR, S100A9, LOX-1) markers, as well as dual CD8-PD-L1 and CD4-FOXP3 staining, were used to quantify immune-cell densities and spatial proximity. Sections were stained using Ventana Benchmark XT (Roche) platforms and visualised with OptiView and UltraView detection kits. Slides were digitised at 20× resolution (NanoZoomer XR, Hamamatsu) and analysed using the HALO software (Indica Labs) for region annotation, cell segmentation, phenotype classification, and spatial analysis. Critically, all immune-cell density quantifications were performed separately within three spatially distinct tissue compartments — tumour, stroma, and parenchyma — enabling compartment-specific characterisation of the immune microenvironment rather than bulk tissue-level estimation.

### 7. Multiplex immunofluorescence — imaging and analysis details

Multiplex immunofluorescence was performed using three panels targeting innate immunity, NK/T cells, and B cells on sequential FFPE sections. Staining was automated on the Bond RX platform (Leica) with the Opal 6-Plex kit (Akoya Biosciences). Slides were imaged on the PhenoImager HT platform at 20× magnification using seven spectral filters. Unmixing and image processing were conducted using inForm (Akoya), followed by quantitative analysis in HALO v3.4 (Indica Labs). Tumour and normal-adjacent tissue regions were manually annotated, and the Highplex FL module in HALO v3.4 was used to identify and phenotype individual cells.

### 8. Genomic and transcriptomic sequencing — detailed protocols

Macrodissection of tumour regions was performed from 1–14 FFPE slides. DNA and RNA were co-extracted using the Quick-DNA/RNA FFPE Miniprep Kit (Zymo Research), and matched germline DNA was extracted from peripheral blood pellets using the DNeasy Blood & Tissue Kit (Qiagen). Whole-exome sequencing libraries were enriched with the xGen Exome Hyb Panel (IDT Technologies) and sequenced on an Illumina NovaSeq platform to achieve mean target coverage of 100× for tumour and 70× for normal tissue. TMB was calculated from coding non-synonymous variants per megabase of tumour DNA, as previously described^2,3^.

Mutational analyses were restricted to a curated panel of 32 genes, selected to match the targeted sequencing panel routinely used by the pathology department: *KRAS, HER2, TP53, CTNNB1, STK11, DDR2, FBXW7, SMAD4, ERBB4, BRAF, FGFR2, FGFR3, PDGFRA, PTEN, MET, TERT, NRAS, PIK3CA, EGFR, KIT, POLE, RET, HRAS, IDH1, KIF5B, ERBB2, ROS1, TTF1, ANTI-CYTOKERATINE, CDKN2A, PDL1, KEAP1*.

Total RNA extracted from FFPE sections was quantified and quality-controlled using Qubit RNA HS (Thermo Fisher) and Agilent ScreenTape. Whole-transcriptome libraries were prepared using the QuantSeq 3ʹ mRNA-Seq Kit (Lexogen) and sequenced on an Illumina NextSeq 2000 to generate 75-bp single-end reads.

### 9. Data infrastructure and reproducibility

Computational pre-processing, biostatistical analyses, and supervised modelling were performed in a controlled containerised environment encompassing fixed operating-system, R, Python, and package versions. Three internally developed open-source packages were used for the analyses: compo.EDA for biostatistical analyses and reporting^4^, compo.tidyML for machine learning analyses and reporting^5^, and ROOFS for robust biomarker feature selection^6^. A continuous integration and deployment workflow was implemented to ensure data-integrity testing, traceability, reproducibility, and centralised reporting of the analyses.

A dedicated companion website was developed, enabling interactive interrogation of biomarker distributions and associations with multiple outcomes, correlation structures, alongside comprehensive statistical and machine learning reports, in support of open science (https://compo.inria.fr/pioneer-website/).

### 10. Preprocessing — imputation strategy evaluation and multicollinearity rationale

Given the expected high degree of multicollinearity within biological data layers — particularly among immune cell subsets quantified by overlapping phenotypic markers, and among correlated biochemical variables — multicollinearity was systematically addressed using iterative variance inflation factor (VIF) analysis^7^. Variables with VIF > 5 were iteratively removed until all remaining variables met acceptable collinearity thresholds (VIF ≤ 5), yielding a final decorrelated dataset of 214 features. All univariable biostatistical and machine learning analyses reported in the main manuscript were performed on this 214-feature decorrelated dataset. Complete results for all 442 profiled biomarkers — including markers removed during multicollinearity reduction — are reported on the companion website (https://compo.inria.fr/pioneer-website/).

Within the machine learning pipelines, missing values were imputed using the median for numeric variables and the mode for categorical variables. Multiple imputation strategies — including k-nearest neighbours and multiple imputation by chained equations — were evaluated but yielded no improvement in predictive performance over simple median/mode imputation, which was therefore retained. Multi-level categorical variables were one-hot encoded. Imputation, encoding, and scaling were embedded within the modelling pipelines to prevent information leakage between training and testing sets.

### 11. Univariable biostatistical analysis — detailed tests

Associations between individual biomarkers and the binary PrR endpoint were assessed using logistic regression and appropriate statistical tests. Numeric variables were analysed with t tests (sample size ≥ 30) or Wilcoxon tests (sample size < 30), and categorical variables with chi-squared tests (sample size ≥ 50) or Fisher’s exact tests (sample size < 50). Area under the receiver operating characteristic curve (AUROC) was computed for each marker.

Associations with PFS were assessed using Cox proportional-hazards regression, complemented by log-rank testing on stratified Kaplan-Meier curves. For each continuous feature, an optimal cutpoint was derived using the maximally selected rank statistic implemented in the surv_cutpoint function from the survminer package, constrained to retain at least 10% of patients in each group. Harrell’s concordance statistic (C-index) was also computed.

To identify associations independent of PD-L1 expression, both logistic and Cox regression models were repeated with adjustment for PD-L1 as a continuous covariate. To account for multiple testing across the large number of biomarkers, adjusted p values (q values) were computed using the Benjamini-Hochberg false discovery rate procedure^8^. These analyses were implemented within the compo.EDA package^4^.

### 12. Optimism-correction — derivation and performances assessment

#### 12.1 Derivation

The modelling workflows, including preprocessing (imputation, scaling), feature selection, model selection, and hyperparameter tuning, were implemented via ROOFS^6^, an internally developed Python package built on the scikit-learn library^9^. ROOFS relies on the resampling-based .632 optimism correction method for rigorous internal validation^10,11^. Briefly, identical modelling pipelines were applied to both the full dataset and each resample. The estimate of a given performance metric *θ* was calculated as a weighted sum of the apparent and out-of-bag performance:

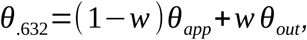

where *θ_app_* is the performance metric computed on the full dataset, and *θ_out_* is the performance metric computed on external samples (patients not included in the resampled dataset), averaged across resamples. In the standard .632 method, the weight *w*=0.632 corresponds to the approximate number of samples included in a bootstrap. Since we used subsampling without replacement to 80% of the original number of patients, we adjusted the weight to w = 0.8 accordingly to reflect the actual proportion of in-bag samples. One hundred subsamples per pipeline were used.

#### 12.2 Assessment of discrimination and calibration

The primary discrimination evaluation metrics were the .632 corrected AUC and Positive Predictive Value (PPV) for prediction of PrR. Secondary metrics were the .632 corrected balanced accuracy, sensitivity, Negative Predictive Value (NPV), and specificity.

To evaluate the calibration of the final model, calibration curves were constructed by plotting mean predicted probabilities against observed event rates. For apparent calibration — predictions generated for all patients using the full model — probabilities were grouped into 10 equal-width bins. For out-of-bag calibration — predictions obtained from out-of-bag samples, with approximately 88 patients per subsample — probabilities were grouped into 7 equal-width bins.

Decision curve analysis was performed to evaluate the clinical utility of the final model in reducing unnecessary treatment in patients unlikely to benefit. Net benefit curves were constructed for the final model and compared with PD-L1 expression alone, the "intervention for all" strategy (intervention being avoidance of treatment), and the "intervention for none" strategy (all patients receive treatment). Net benefit was calculated across 100 evenly spaced probability thresholds *p_t_ ∈* (0, 1) according to:

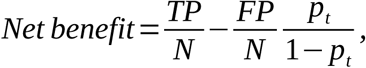

where *TP* denotes the number of true positives, *FP* the number of false positives, and *N* the total sample size.

### 13. Feature selection benchmarking — stability metric and full method enumeration

#### 13.1. Stability metric

Thirty-six feature selection (FS) methods were benchmarked for performance and stability^12^. The Nogueira stability index is defined as:

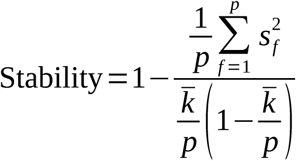

where p is the total number of features in the dataset, 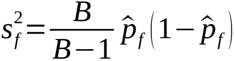 is the unbiased sample variance of 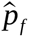, the frequency of selection of the feature *f* over the *B* subsamples and 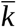 is the average number of selected features in a subsampled dataset (thus 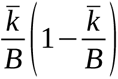 is the expected variance of the frequency of selection for a uniformly randomly selected feature).

#### 13.2. Methods evaluated

The FS methods included embedded methods: Least Absolute Shrinkage and Selection Operator, LASSO^13^, stability selection^14^, bootstrap LASSO^15^, adaptive Lasso^16^, relaxed Lasso^17^, sparse group Lasso with correlation and Minkowski distances^18^, Stabl LASSO and adaptive Lasso^19^, and RENT^20^; filter methods: Fisher score^21^, trace ratio^22^, Conditional Infomax Feature Extraction (CIFE)^23^, Conditional Mutual Information (CMIM)^24^, Double Input Symmetrical Relevance (DISR) filter^25^, Interaction CAPping (ICAP)^26^, Joint Mutual Information (JMI)^27^, Mutual Information Maximization (MIM)^28^, ReliefF^29^, minimum Redundancy Maximum Relevance (mRMR)^30^, F-score^21^, Gini index^21^, hierarchical clustering on the correlation matrix (selecting the feature with the highest correlation with the outcome within each cluster), and t-score^21^; and wrapper methods: recursive feature elimination using either logistic regression or random forest^31^, recursive forward selection with random forest, and Shapicant^32^. For filter methods that required a predefined number of features, it was fixed to 15. We additionally employed univariable PFS-Cox or PrR-logistic regression (controlled by PD-L1) or t-test-based filtering with an adjusted p-value threshold of 0.05. All the methods were grouped in ROOFS, allowing unified benchmarking^6^.

### 14. Predictive modelling — hyperparameters

Ten machine learning models, including logistic regression, random forests, (extreme) gradient boosting, and multi-layer perceptrons, were evaluated with the scikit-learn-based ROOFS package^6^. Hyperparameter tuning yielded negligible gains and was not retained in the final models. For tree-based models in the main analysis, maximum tree depth was limited to 2, the minimum number of patients per leaf node was set to 40, and the number of estimators was set to 200. Logistic regression was fitted without regularisation.

### 15. PFS stratification analysis — design and rationale

The final model was also evaluated for its ability to stratify PFS. A stratified train-test split (2/3– 1/3) was generated after k-means clustering (k = 5) on known prognostic variables (PD-L1 expression at the 1% threshold, histology, performance status, and sex), with cluster used as the stratification variable. This stratification ensured that the prognostic structure of the cohort was preserved across the training and test sets, mitigating risk of imbalance for low-frequency clinico-pathological subgroups. The fitted model generated a predicted probability of PrR, which was then used to stratify PFS. The cutpoint for this score was defined in the training set as the value that optimally separated Kaplan-Meier PFS curves using surv_cutpoint (minprop = 0.1).

### 16. Explainability analyses — SHAP and decision tree parameters

To interpret the contribution of individual markers to PrR prediction, SHapley Additive exPlanation (SHAP) analysis was conducted in both the feature and patient dimensions using the kernelshap^33^ and shapviz^34,35^ R packages. For patient-level analyses, the model was trained on the training set described above, and predictions with SHAP contributions were generated in the test set.

In addition, decision trees were built on the signature variables using DecisionTreeClassifier (max_depth = 4, min_samples_leaf = 10, ccp_alpha = 0.01) from scikit-learn^9^.

## Supplementary figures and tables

**Supplementary Table 1: Dictionary of biomarkers Legend.** Comprehensive data dictionary for all biomarkers considered in this study. The Excel file is provided in Supplementary Table 1, including variable names, technology, units, and whether kept or not at different stages of the analysis.

**Supplementary Table 2:**
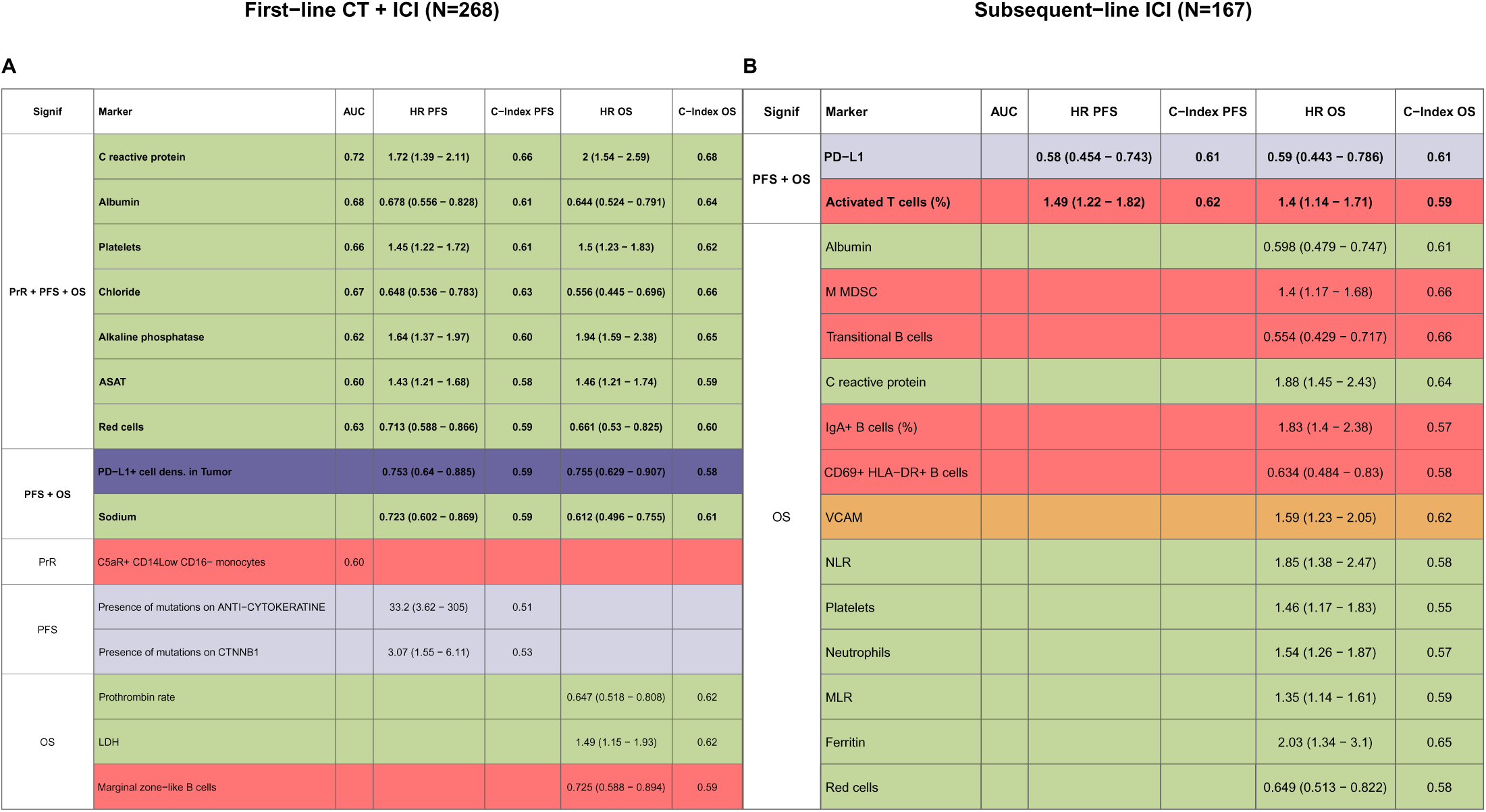
Statistical summary across all lines. **Legend.** Summary of significant biomarkers across treatment lines, showing discrimination for classification tasks (AUC) and prognostic performance for time-to-event tasks (hazard ratio with 95% confidence interval and C-index for PFS and OS). Results are reported separately for (A) first-line chemo-immunotherapy and (B)subsequent-line ICI cohorts.

**Supplementary Table 3:** Comparison with literature. **Legend.** Benchmark of PIONeeR against recent multimodal immuno-oncology studies. DCB: Durable Control Benefit, similar to primary resistance; EP: Early Progression (progression ≤ 3 months); ORR: Objective Response Rate; PFS: Progression-Free Survival; OS: Overall Survival; ICI: Immune-Checkpoint Inhibition; CT: Chemotherapy. *: C-index from a Cox model re-fitted on the validation cohort. **: median of performances on validation sets

| Study | Cohort size | Data modalities | DCB | EP | ORR | PFS | OS | External validation | ICI | ICI+CT |
| --- | --- | --- | --- | --- | --- | --- | --- | --- | --- | --- |
| Barlesi (PIONeeR) 2026 | 439<br>IO+CT : 269<br>IO : 170 | Clinical<br>Blood tests<br>Multiplex IHC + IF<br>Immuno-phenotyping<br>Vasculo-phenotyping<br>Genomic | 0.73 ± 0.05 | 0.78 ± 0.03 | 0.66 ± 0.04 | 0.69 ± 0.03 | 0.69 ± 0.03 | — | ✓ | ✓ |
| Captier Nat Comm 2025 | 317<br>(80 all modalities) | Clinical<br>Radiomic<br>Pathomic<br>Transcriptomic | 0.67 ± 0.03 |  |  | ~0.63 ± 0.02 | D : 0.75 ± 0.01 | — | ✓ | ✓ |
| Yoo (SCORPIO) Nat Med 2025 | D : 2,035; V : 7,710 | Clinical<br>Blood tests | D : 0.714<br>V : 0.63** |  |  |  | V : 0.71** | ✓ | ✓ | ✓ |
| Rakaee (Deep-IO) JAMA Oncol 2025 | D : 614 ; V : 344 | Pathologic |  |  | D : 0.75±0.05<br>V : 0.66±0.03 | V : 0.65* | V : 0.64* | ✓ | ✓ | — |
| Chang (LORIS) Nat Cancer 2024 | D : 324 ; V : 612 | Clinical<br>Blood tests |  |  | D : 0.74 ± 0.03<br>V : 0.725** | HR=2.6<br>(95%CI 2.2–3.0) | HR=3.2<br>(95%CI 2.6–3.9) | ✓ | ✓ | — |
| Hong Nat Comm 2023 | D : 1133<br>IO+CT : 407<br>IO : 726<br>V : 89<br>IO+CT : 49<br>IO : 40 | Clinical<br>Genomic |  | IO + CT<br>D : 0.66<br>IO<br>D : 0.73, V : 0.73 |  |  |  | ✓ | ✓ | ✓ |
| Vanguri (DyAM) Nat Cancer 2022 | 247<br>(81 all modalities) | Radiologic<br>Pathologic<br>Genomic |  |  | D : 0.80 ± 0.03 |  |  | — | ✓ | — |

**Supplementary Figure 1:**
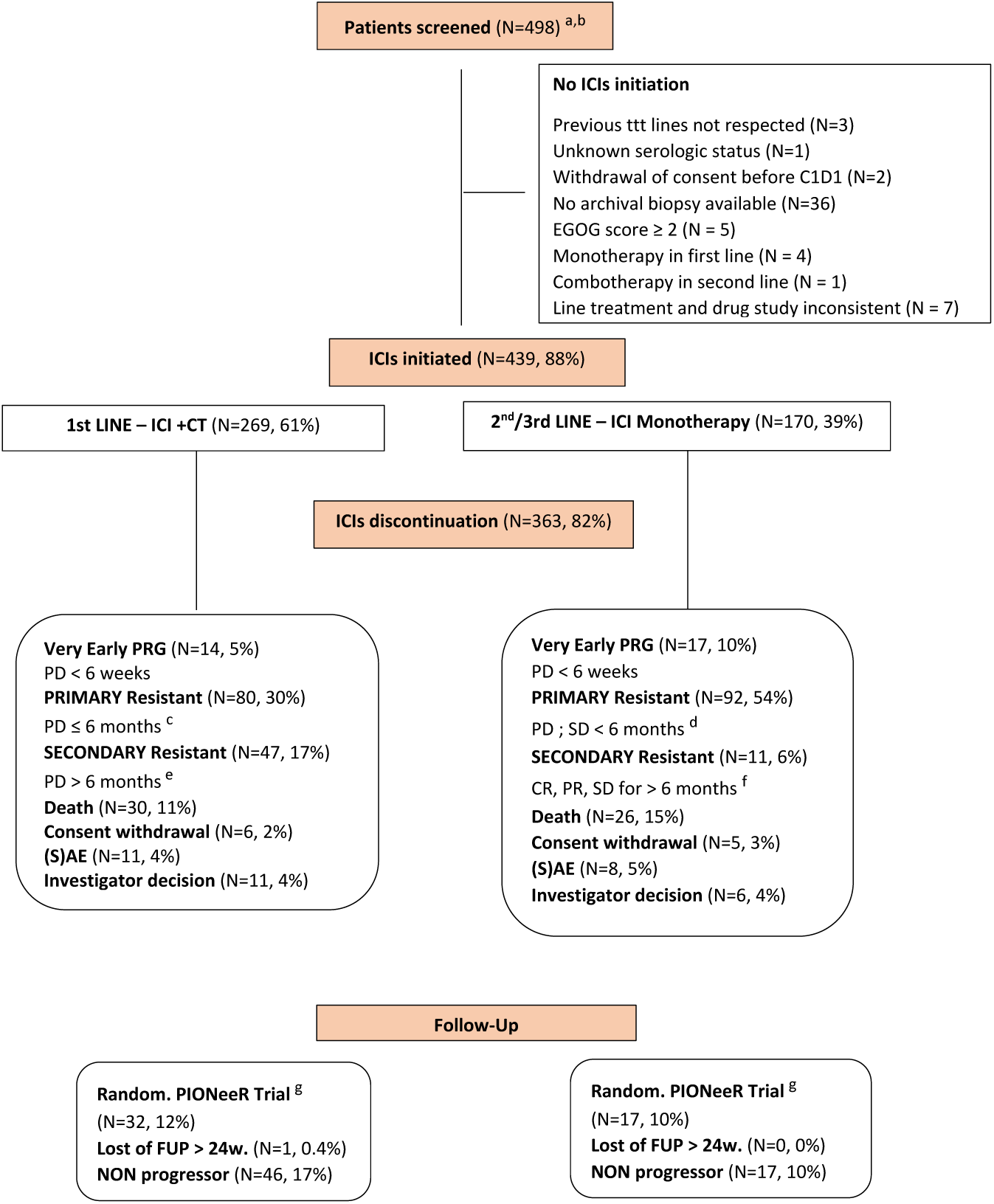
Flowchart. **Legend.** CONSORT-style flow diagram of patient screening, eligibility, treatment-line assignment (first-line ICI+CT versus subsequent-line ICI), treatment discontinuation, and follow-up outcome categories (very early progression, primary resistance, secondary resis-tance, non-progressor). (a)Advanced NSCLC patients registered for PD1/L1 ICI-based treatment. (b) Informed consent signed. (c) According to SITC guidelines (Rivzi, 2023 + Tawbi 2023), Best Response according to RECIST v1.1 with a treatment exposure ≥ 6-8 weeks. (d) a treatment exposure ≥ 6 weeks. (e) a treatment exposure > 6 months. (f) a treatment exposure ≥ 6 months. (g) NCT03833440. (h) Eligible /Reaching biopsy time, N=349

**Supplementary Figure 2:**
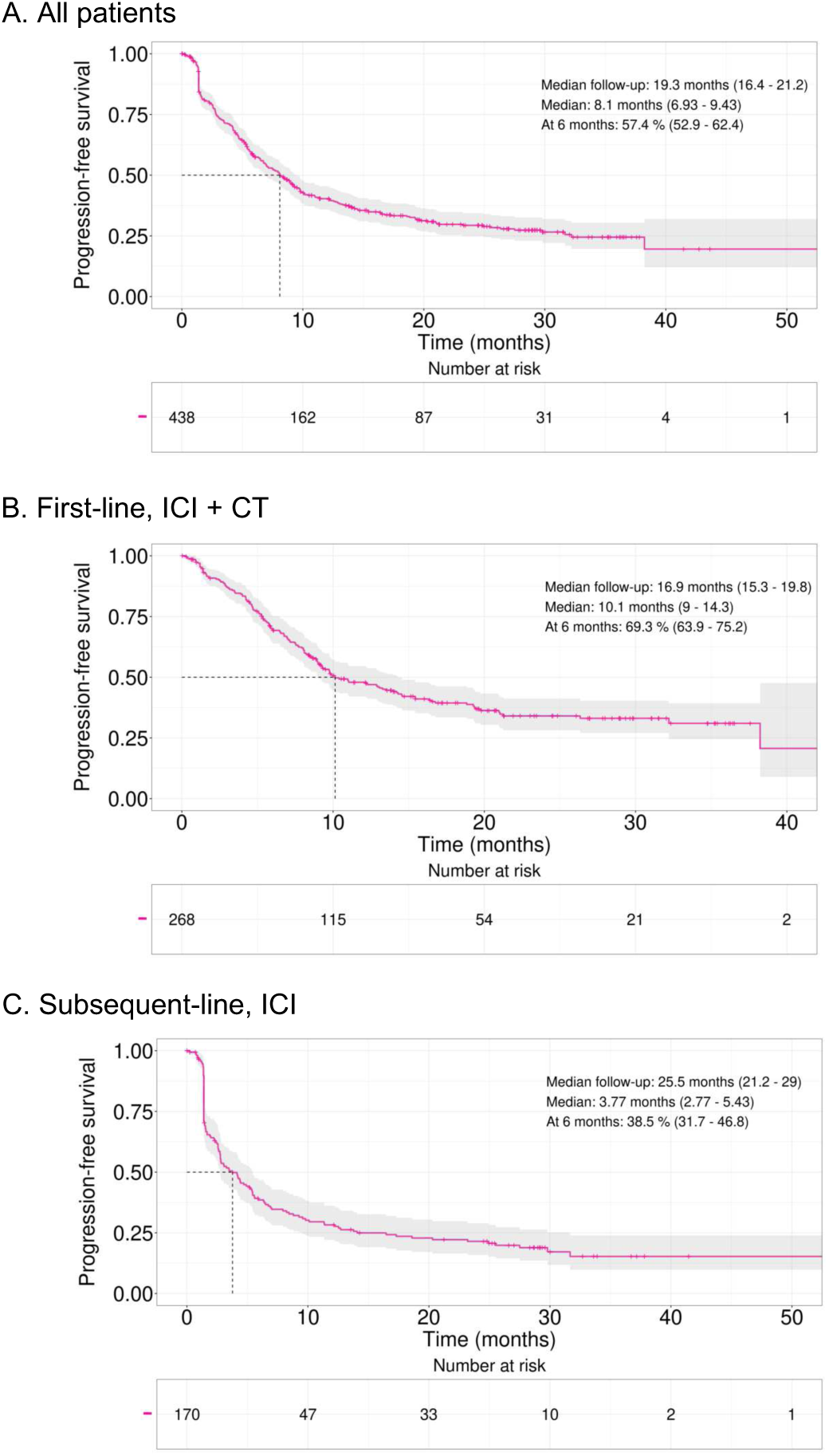
Progression-free survival. **Legend.** Kaplan-Meier estimates of progression-free survival in the full population and stratified by treatment setting: (A) all patients, (B) first-line ICI+chemotherapy, and (C) subsequent-line ICI monotherapy.

**Supplementary Figure 3:**
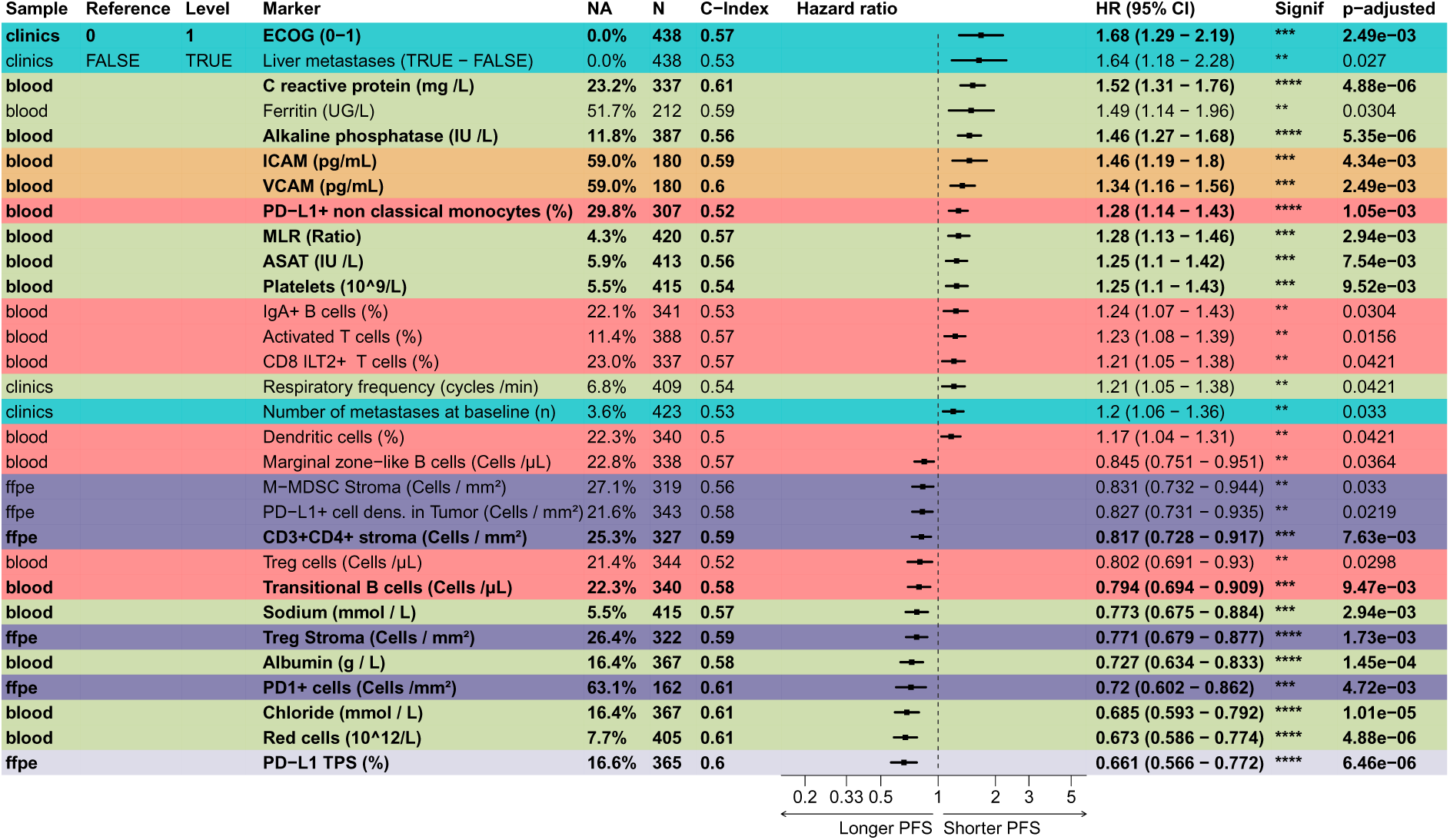
Forest plot for progression-free survival. **Legend.** Univariable forest plot of candidate biomarkers and clinical covariates for progression-free survival (BH-filtered PFS-Cox regression q ¡ 0.05). For each feature, the plot reports reference level (when categorical), hazard ratio with 95% confidence interval, C-index, and adjusted significance, enabling cross-domain comparison of prognostic strength. Bold biomarkers: q < 0.01, selected for the signature as per the best feature-selection method.

**Supplementary Figure 4:**
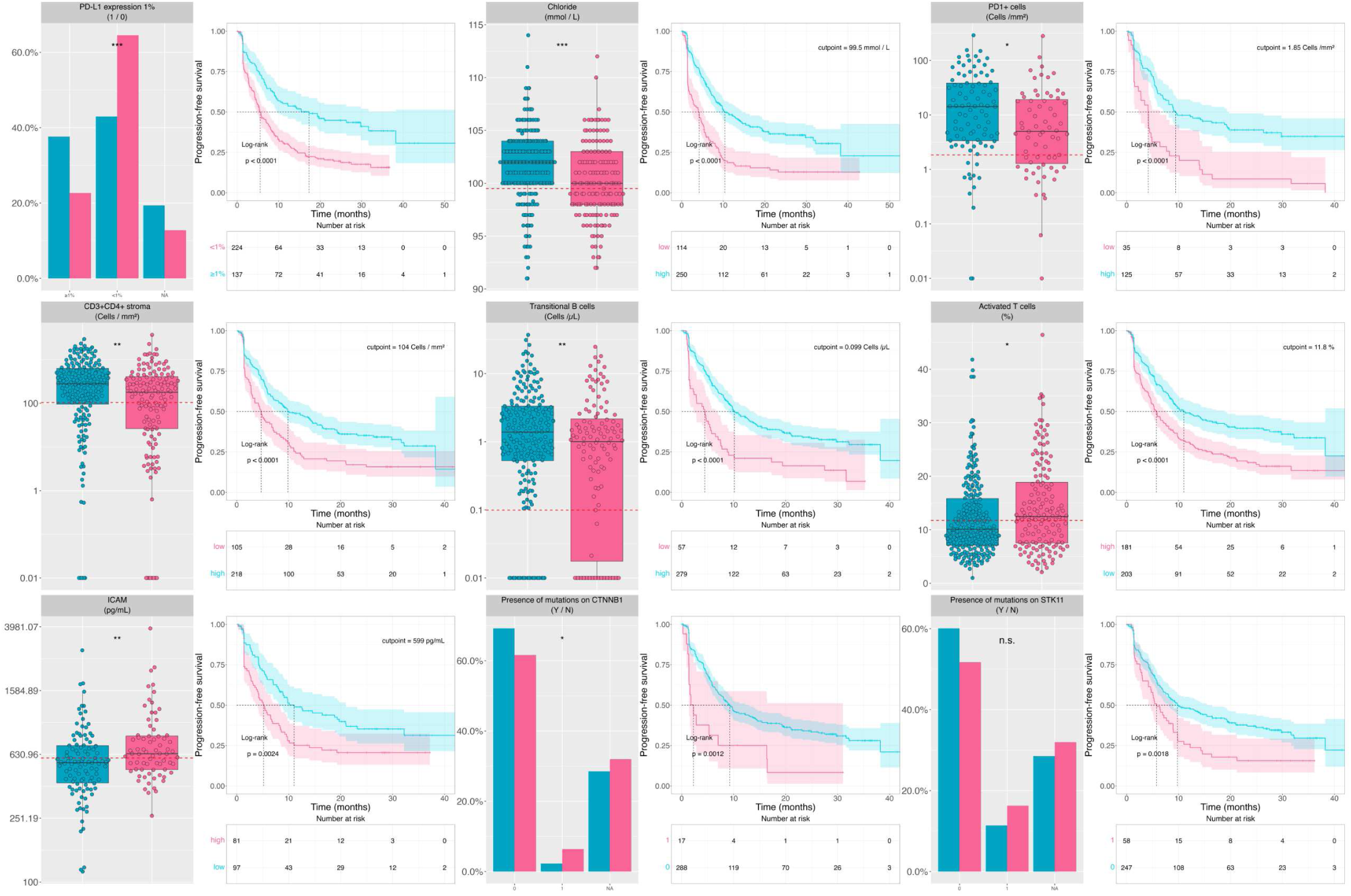
Additional PrR boxplots and PFS Kaplan-Meier plots. **Legend.** Joint visualization of additional significant biomarkers in univariable analyses.

**Supplementary Figure 5:**
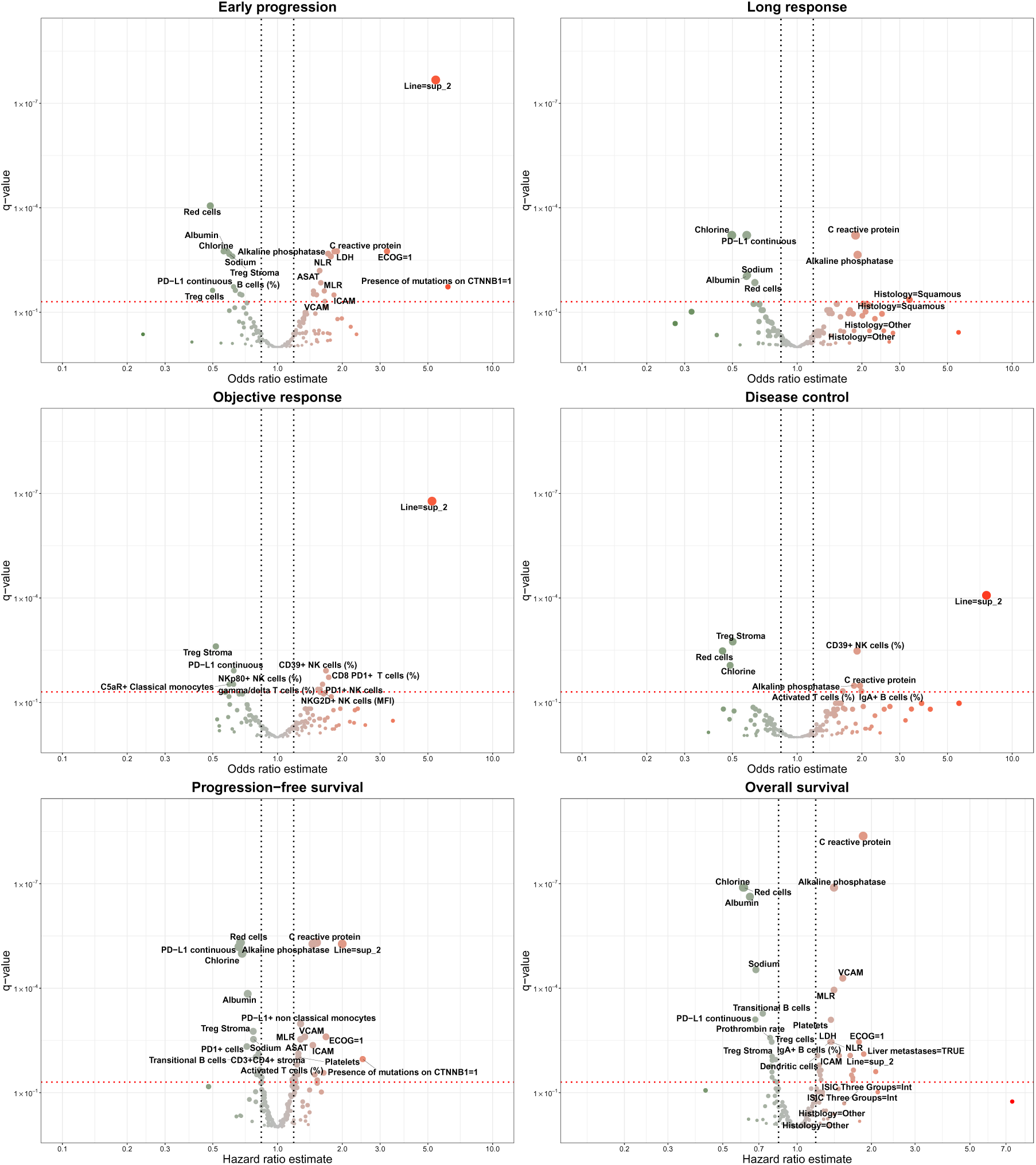
Volcano plots of secondary endpoints. **Legend.** Volcano plots of association analyses across endpoints: primary resistance, early progression, overall response, disease control, progression-free survival, and overall survival. Effect-size estimates (odds ratio or hazard ratio) are shown against multiple-testing-adjusted q-values, with direction and strength of associations across biomarkers and clinical variables.

**Supplementary Figure 6:**
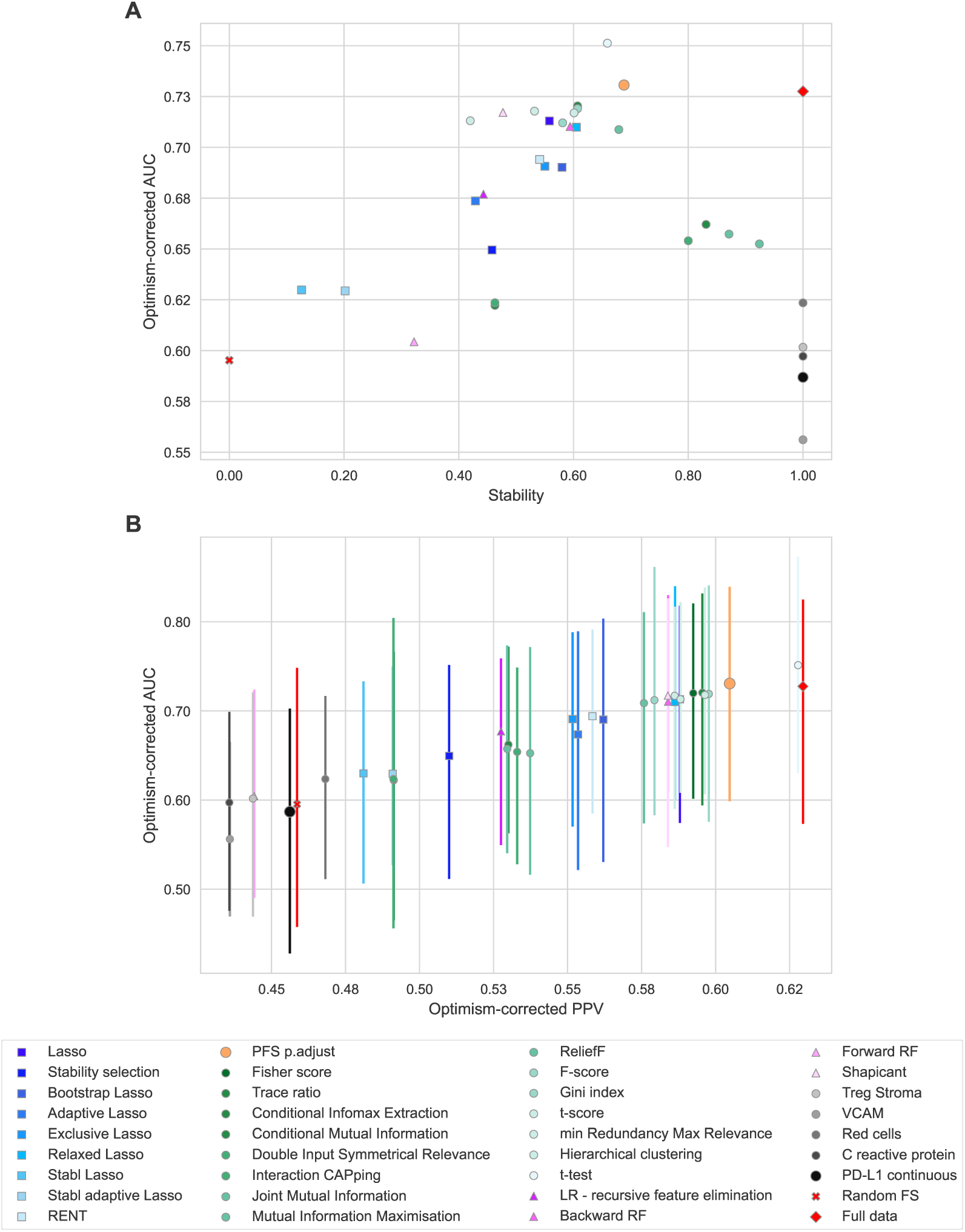
Metrics versus feature-selection stability. **Legend.** Comparison of feature-selection (FS) strategies using stability and predictive performance metrics. Panels show how optimism-corrected model performance (AUROC/PPV) varies with feature-selection reproducibility across bootstrap, lasso-based, filter, and embedded selection methods. PFS p.adjust: selected FS method at the pareto front of stability and predictive performance, consisting of Benjamini-Hochberg adjusted q-value ¡ 0.01 for Cox regression on PFS. The t-test filter – with similar stability and slightly higher AUROC and PPV was discarded for parsimony (mean number of variables 43.4 versus 12.5)

**Supplementary Figure 7:**
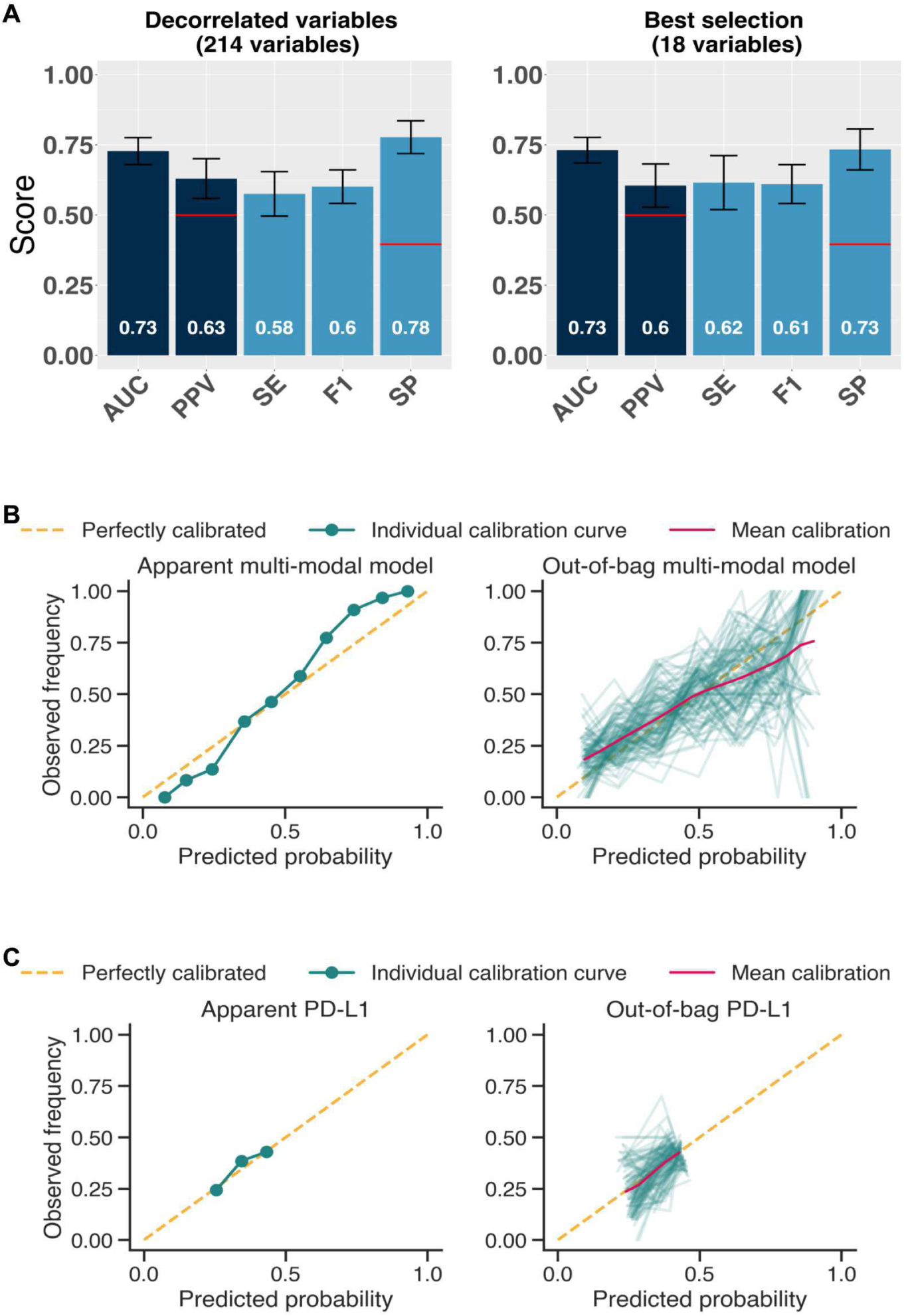
Discrimination and calibration metrics of the multimodal machine learning model. **Legend.** Model discrimination and calibration diagnostics across prediction tasks. A. Discrimination metrics. B. Calibration of the minimal multimodal model. The left panel shows the apparent calibration curve obtained on the full dataset by comparing predicted probabilities with observed event frequencies across probability bins; the dashed line indicates perfect calibration. The right panel shows resampling out-of-bag calibration, where each thin green line represents one out-of-bag calibration curve and the magenta line represents the mean calibration across resamples. C. Calibration of the PD-L1 model, built as a logistic regression on PD-L1 tumor proportion score. AUC, area under the receiver operating characteristic curve; PPV, positive predictive value; SE, sensitivity; SP, specificity; F1, F1 score

**Supplementary Figure 8:**
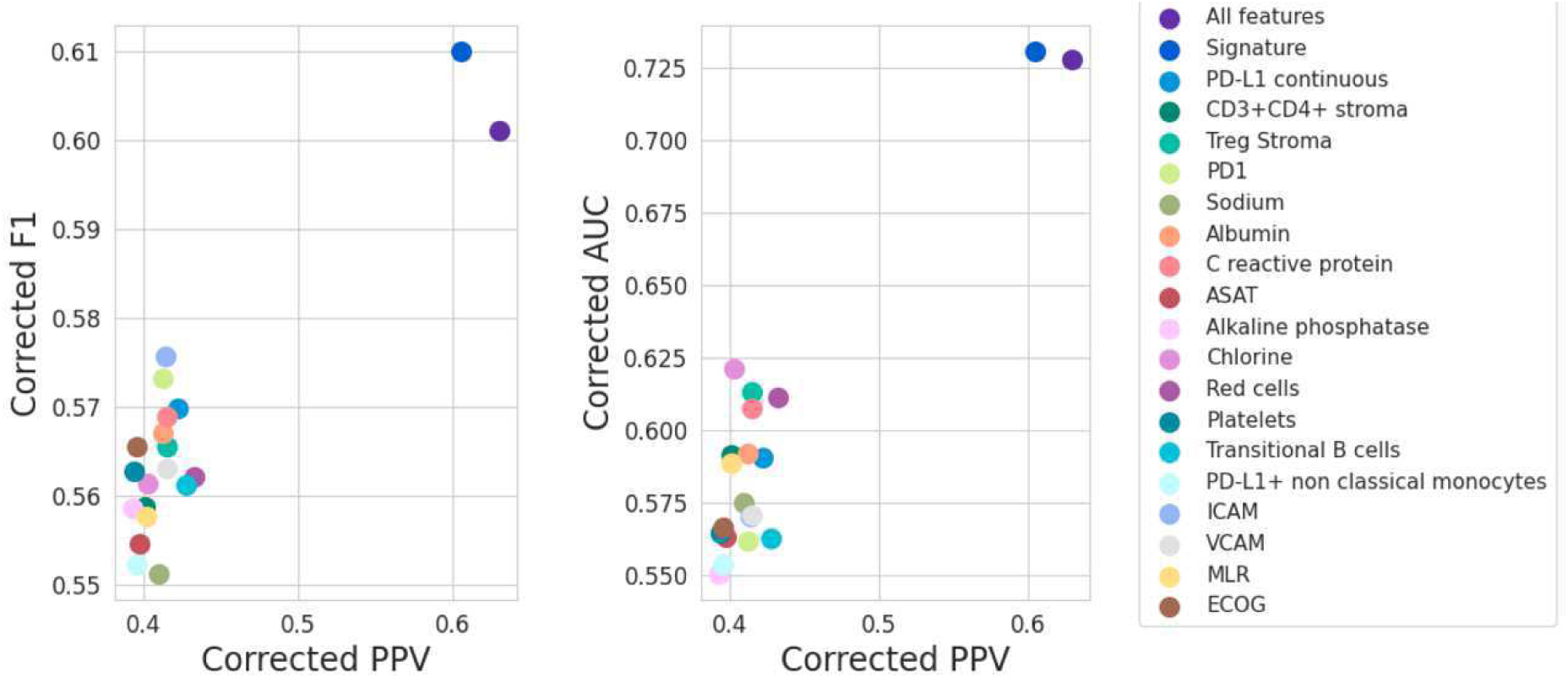
Metrics for individual biomarkers. **Legend.** Performance summary of single biomarkers versus the full (all – decorrelated – features) and minimal multimodal machine learning model. AUC, area under the receiver operating characteristic curve; PPV, positive predictive value; F1, F1 score

**Supplementary Figure 9:**
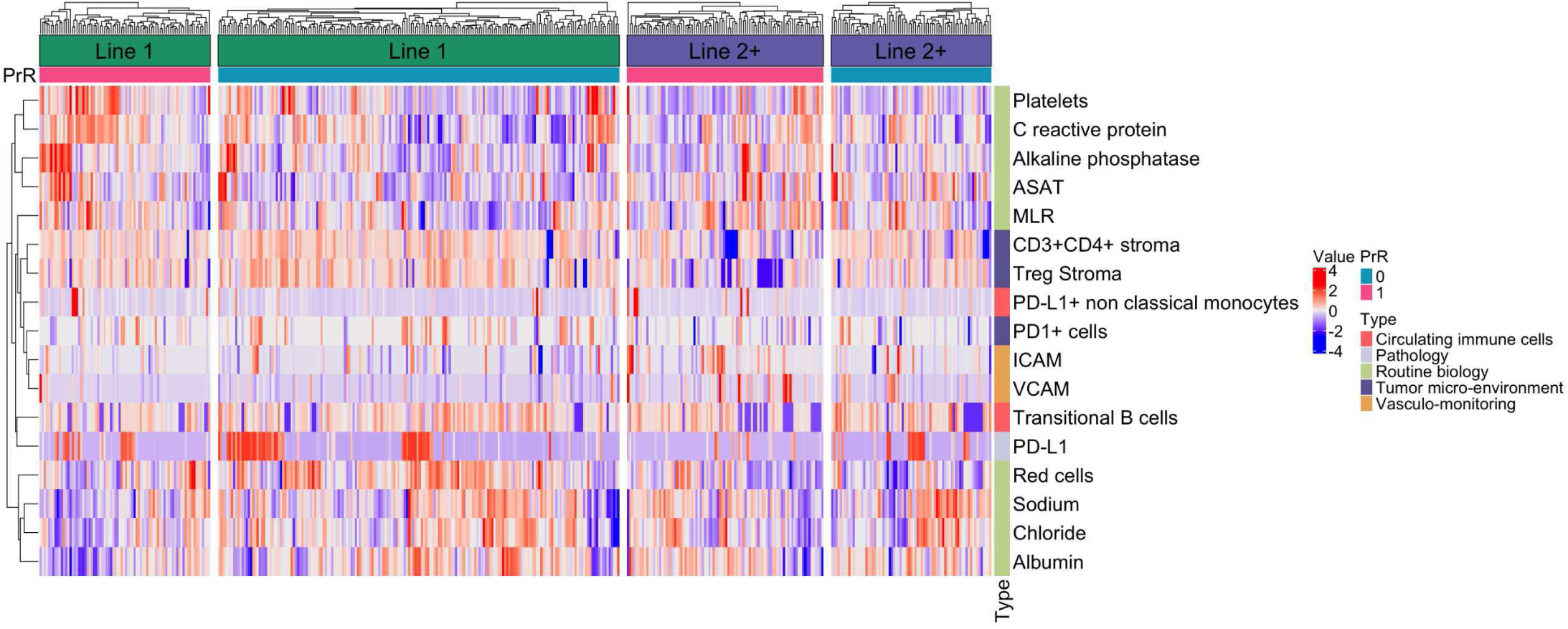
Hierarchical clustering of patients on the 18-feature multimodal signature. **Legend.** Heatmap of scaled signature variable values (rows) across evaluable patients (columns), grouped by treatment line (Line 1: first-line ICI + chemotherapy; Line 2+: subsequent-line ICI monotherapy) and primary resistance status (n = 435, PrR = 1, pink; PrR = 0, teal). Within each stratum, patients are ordered by hierarchical clustering (Euclidean distance, complete linkage); signature variables are likewise hierarchically clustered (left dendrogram). PrR: primary resistance; ICI: immune checkpoint inhibitor; CT: chemotherapy.

## REFERENCES

1. Hendriks, L. E. et al. Non-oncogene-addicted metastatic non-small-cell lung cancer: ESMO Clinical Practice Guideline for diagnosis, treatment and follow-up. Ann Oncol 34, 358–376 (2023).

2. Wang, Y. et al. Immunotherapy for advanced-stage squamous cell lung cancer: the state of the art and outstanding questions. Nat Rev Clin Oncol 22, 200–214 (2025).

3. Gandhi, L. et al. Pembrolizumab plus Chemotherapy in Metastatic Non-Small-Cell Lung Cancer. N. Engl. J. Med. 378, 2078–2092 (2018).

4. Herbst, R. S. et al. Pembrolizumab versus docetaxel for previously treated, PD-L1-positive, advanced non-small-cell lung cancer (KEYNOTE-010): a randomised controlled trial. The Lancet 387, 1540–1550 (2016).

5. Reck, M. et al. Pembrolizumab versus Chemotherapy for PD-L1–Positive Non– Small-Cell Lung Cancer. N. Engl. J. Med. 375, 1823–1833 (2016).

6. Mok, T. S. K. et al. Associations of tissue tumor mutational burden and mutational status with clinical outcomes in KEYNOTE-042: pembrolizumab versus chemotherapy for advanced PD-L1-positive NSCLC. Ann Oncol 34, 377–388 (2023).

7. Mok, T. S. K. et al. Pembrolizumab versus chemotherapy for previously untreated, PD-L1-expressing, locally advanced or metastatic non-small-cell lung cancer (KEYNOTE-042): a randomised, open-label, controlled, phase 3 trial. Lancet 393, 1819–1830 (2019).

8. Reck, M. et al. Nivolumab plus ipilimumab versus chemotherapy as first-line treatment in advanced non-small-cell lung cancer with high tumour mutational burden: patient-reported outcomes results from the randomised, open-label, phase III CheckMate 227 trial. Eur J Cancer 116, 137–147 (2019).

9. Ricciuti, B. et al. Association of High Tumor Mutation Burden in Non-Small Cell Lung Cancers With Increased Immune Infiltration and Improved Clinical Outcomes of PD-L1 Blockade Across PD-L1 Expression Levels. JAMA Oncol 8, 1160–1168 (2022).

10. Cristescu, R. et al. Pan-tumor genomic biomarkers for PD-1 checkpoint blockade-based immunotherapy. Science 362, (2018).

11. Yarchoan, M., Hopkins, A. & Jaffee, E. M. Tumor Mutational Burden and Response Rate to PD-1 Inhibition. N Engl J Med 377, 2500–2501 (2017).

12. Hellmann, M. D. et al. Genomic Features of Response to Combination Immunotherapy in Patients with Advanced Non-Small-Cell Lung Cancer. Cancer Cell 33, 843–852 e4 (2018).

13. Skoulidis, F. et al. STK11/LKB1 Mutations and PD-1 Inhibitor Resistance in KRAS-Mutant Lung Adenocarcinoma. Cancer Discov. 8, 822–835 (2018).

14. Schoenfeld, A. J. et al. The Genomic Landscape of SMARCA4 Alterations and Associations with Outcomes in Patients with Lung Cancer. Clin. Cancer Res. 26, 5701–5708 (2020).

15. Skoulidis, F. et al. CTLA4 blockade abrogates KEAP1/STK11-related resistance to PD-(L)1 inhibitors. Nature 635, 462–471 (2024).

16. Tumeh,P. C. et al. PD-1 blockade induces responses by inhibiting adaptive immune resistance. Nature 515, 568–571 (2014).

17. Cabrita, R. et al. Tertiary lymphoid structures improve immunotherapy and survival in melanoma. Nature 577, 561–565 (2020).

18. Ayers, M. et al. IFN-gamma-related mRNA profile predicts clinical response to PD-1 blockade. J Clin Invest 127, 2930–2940 (2017).

19. Rakaee, M. et al. Deep Learning Model for Predicting Immunotherapy Response in Advanced Non-Small Cell Lung Cancer. JAMA Oncol 11, 109–118 (2025).

20. Mezquita, L. et al. Association of the Lung Immune Prognostic Index With Immune Checkpoint Inhibitor Outcomes in Patients With Advanced Non–Small Cell Lung Cancer. JAMA Oncol. 4, 351–357 (2018).

21. Lee, J. S. & Ruppin, E. Multiomics Prediction of Response Rates to Therapies to Inhibit Programmed Cell Death 1 and Programmed Cell Death 1 Ligand 1. JAMA Oncol 5, 1614–1618 (2019).

22. Captier, N. et al. Integration of clinical, pathological, radiological, and transcriptomic data improves prediction for first-line immunotherapy outcome in metastatic non-small cell lung cancer. Nat. Commun. 16, 614 (2025).

23. Vanguri, R. S. et al. Multimodal integration of radiology, pathology and genomics for prediction of response to PD-(L)1 blockade in patients with non-small cell lung cancer. Nat Cancer 3, 1151–1164 (2022).

24. Chang, T.-G. et al. LORIS robustly predicts patient outcomes with immune checkpoint blockade therapy using common clinical, pathologic and genomic features. *Nat*. Cancer 5, 1158–1175 (2024).

25. Yoo, S.-K. et al. Prediction of checkpoint inhibitor immunotherapy efficacy for cancer using routine blood tests and clinical data. Nat. Med. 31, 869–880 (2025).

26. Atkins, M. B. et al. Society for Immunotherapy of Cancer (SITC) consensus definitions for resistance to combinations of immune checkpoint inhibitors with targeted therapies. J Immunother Cancer 11, (2023).

27. Rizvi, N. et al. Society for Immunotherapy of Cancer (SITC) consensus definitions for resistance to combinations of immune checkpoint inhibitors with chemotherapy. J Immunother Cancer 11, (2023).

28. Eisenhauer, E. A. et al. New response evaluation criteria in solid tumours: Revised RECIST guideline (version 1.1). Eur. J. Cancer 45, 228–247 (2009).

29. Vallier, L. et al. Increasing the sensitivity of the human microvesicle tissue factor activity assay. Thromb Res 182, 64–74 (2019).

30. Chalmers, Z. R. et al. Analysis of 100,000 human cancer genomes reveals the landscape of tumor mutational burden. Genome Med 9, 34 (2017).

31. Scholler, N. et al. Tumor immune contexture is a determinant of anti-CD19 CAR T cell efficacy in large B cell lymphoma. Nat. Med. 28, 1872–1882 (2022).

32. Nguyen Phuong, L., Vaglio, A. & Benzekry, S. compo.EDA: Data Analysis for Oncology. https://gitlab.inria.fr/compo/compo.eda. (2025).

33. Vaglio, A. & Benzekry, S. compo.tidyML. https://gitlab.inria.fr/compo/compo.tidyml. (2025).

34. Bakhmach, A. et al. ROOFS: RObust biOmarker Feature Selection. Preprint at 10.48550/arXiv.2601.05151 (2026).

35. Gareth, G., Witten, D., Hastie, T. & Tibshirani, R. An Introduction to Statistical Learning: With Applications in R. (Springer US, 2021).

36. Yekutieli, D. & Benjamini, Y. Resampling-based false discovery rate controlling multiple test procedures for correlated test statistics. J. Stat. Plan. Inference 82, 171–196 (1999).

37. Efron, B. & Tibshirani, R. Improvements on Cross-Validation: The 632+ Bootstrap Method. J. Am. Stat. Assoc. 92, 548–560 (1997).

38. Collins, G. S. et al. Evaluation of clinical prediction models (part 1): from development to external validation. BMJ 384, e074819 (2024).

39. Nogueira, S. On the Stability of Feature Selection Algorithms. J. Mach. Learn. Res. 18, 1–54 (2018).

40. Pedregosa, F. et al. Scikit-learn: Machine Learning in Python. J Mach Learn Res 12, 2825–2830 (2011).

41. Mayer, M. & Watson, D. kernelshap: Kernel SHAP. (2022).

42. Mayer, M. shapviz: SHAP Visualizations. (2022).

43. Lundberg, S. M. & Lee, S.-I. A unified approach to interpreting model predictions. in Proceedings of the 31st International Conference on Neural Information Processing Systems 4768–4777 (Curran Associates Inc., Red Hook, NY, USA, 2017).

44. Boussena, M. et al. Multimodality Stacking with Blockwise missing values and application to the PIONeeR biomarkers study for prediction of resistance to immunotherapy. Preprint at 10.48550/arXiv.2605.25050 (2026).

45. Principe, D. R., Chiec, L., Mohindra, N. A. & Munshi, H. G. Regulatory T-Cells as an Emerging Barrier to Immune Checkpoint Inhibition in Lung Cancer. Front. Oncol. 11, (2021).

46. Zou, W. Immunosuppressive networks in the tumour environment and their therapeutic relevance. Nat. Rev. Cancer 5, 263–274 (2005).

47. Rahal, Z., El Darzi, R., Moghaddam, S. J., Cascone, T. & Kadara, H. Tumour and microenvironment crosstalk in NSCLC progression and response to therapy. Nat. Rev. Clin. Oncol. 22, 463–482 (2025).

48. Taylor, M. H. et al. Phase I dose escalation study of IO-108, an anti-LILRB2 antibody, in patients with advanced solid tumors. J. Immunother. Cancer 12, (2024).

49. Riegler, J. et al. VCAM-1 Density and Tumor Perfusion Predict T-cell Infiltration and Treatment Response in Preclinical Models. Neoplasia 21, 1036–1050 (2019).

50. Hong, L. et al. Efficacy and clinicogenomic correlates of response to immune checkpoint inhibitors alone or with chemotherapy in non-small cell lung cancer. Nat. Commun. 14, 695 (2023).

51. Saad, M. B. et al. Machine-learning driven strategies for adapting immunotherapy in metastatic NSCLC. Nat. Commun. 16, 6828 (2025).

52. Collins, G. S. et al. TRIPOD+AI statement: updated guidance for reporting clinical prediction models that use regression or machine learning methods. BMJ 385, e078378 (2024).

## References

1. Eisenhauer, E. A., Therasse, P., … Verweij, J. New response evaluation criteria in solid tumours: Revised RECIST guideline (version 1.1). European Journal of Cancer 45, 228– 247 (2009).

2. Hellmann, M. D., Nathanson, T., … Wolchok, J. D. Genomic Features of Response to Combination Immunotherapy in Patients with Advanced Non-Small-Cell Lung Cancer. Cancer Cell 33, 843–852 e4 (2018).

3. Chalmers, Z. R., Connelly, C. F., … Frampton, G. M. Analysis of 100,000 human cancer genomes reveals the landscape of tumor mutational burden. Genome Med 9, 34 (2017).

4. Nguyen Phuong, L., Vaglio, A. & Benzekry, S. compo.EDA: Data Analysis for Oncology. https://gitlab.inria.fr/compo/compo.eda. (2025).

5. Vaglio, A. & Benzekry, S. compo.tidyML. https://gitlab.inria.fr/compo/compo.tidyml. (2025).

6. Bakhmach, A., Dufossé, P., … Benzekry, S. ROOFS: RObust biOmarker Feature Selection. (2026) doi:10.48550/arXiv.2601.05151.

7. Gareth, G., Witten, D., Hastie, T. & Tibshirani, R. An Introduction to Statistical Learning: with Applications in R. (Springer US, 2021).

8. Yekutieli, D. & Benjamini, Y. Resampling-based false discovery rate controlling multiple test procedures for correlated test statistics. Journal of Statistical Planning and Inference 82, 171–196 (1999).

9. Pedregosa, F., Varoquaux, G., … Duchesnay, E. Scikit-learn: Machine Learning in Python. J. Mach. Learn. Res. 12, 2825–2830 (2011).

10. Efron, B. & Tibshirani, R. Improvements on Cross-Validation: The 632+ Bootstrap Method. Journal of the American Statistical Association 92, 548–560 (1997).

11. Collins, G. S., Dhiman, P., … Riley, R. D. Evaluation of clinical prediction models (part 1): from development to external validation. BMJ 384, e074819 (2024).

12. Nogueira, S. On the Stability of Feature Selection Algorithms. Journal of Machine Learning Research 18, 1–54 (2018).

13. Tibshirani, R. Regression Shrinkage and Selection via the Lasso. Journal of the Royal Statistical Society. Series B (Methodological) 58, 267–288 (1996).

14. Meinshausen, N. & Bühlmann, P. Stability selection. Journal of the Royal Statistical Society: Series B (Statistical Methodology) 72, 417–473 (2010).

15. Bach, F. R. Bolasso: model consistent Lasso estimation through the bootstrap. in Proceedings of the 25th international conference on Machine learning 33–40 (Association for Computing Machinery, 2008). doi:10.1145/1390156.1390161.

16. Zou, H. The Adaptive Lasso and Its Oracle Properties. Journal of the American Statistical Association 101, 1418–1429 (2006).

17. Meinshausen, N. Relaxed Lasso. Computational Statistics & Data Analysis 52, 374–393 (2007).

18. Simon, N., Friedman, J., Hastie, T. & Tibshirani, R. A Sparse-Group Lasso. Journal of Computational and Graphical Statistics 22, 231–245 (2013).

19. Hédou, J., Marić, I., … Gaudillière, B. Discovery of sparse, reliable omic biomarkers with Stabl. Nature Biotechnology 42, 1581–1593 (2024).

20. Jenul, A., Schrunner, S., … Tomic, O. RENT -- Repeated Elastic Net Technique for Feature Selection. in IEEE Access, *vol.* 9, *pp. 152333-152346*, 2021 9, 152333–152346 (2020).

21. Li, J., Cheng, K., … Liu, H. Feature Selection: A Data Perspective. ACM Computing Surveys 50, 1–45 (2017).

22. Nie, F., Xiang, S., Jia, Y., Zhang, C. & Yan, S. Trace Ratio Criterion for Feature Selection. in inproceedings (eds Fox, D. & Gomes, C. P.) 671–676 ({AAAI} Press, 2008).

23. Lin, D. & Tang, X. Conditional Infomax Learning: An Integrated Framework for Feature Extraction and Fusion. in (eds Leonardis, A., Bischof, H. & Pinz, A.) 68–82 (Springer Berlin Heidelberg, 2006).

24. Fleuret, F. Fast Binary Feature Selection with Conditional Mutual Information. J. Mach. Learn. Res. 5, 1531–1555 (2004).

25. Meyer, P. E. & Bontempi, G. On the Use of Variable Complementarity for Feature Selection in Cancer Classification. in (eds Rothlauf, F. et al.) 91–102 (Springer Berlin Heidelberg, 2006).

26. Jakulin, A. Machine learning based on attribute interactions. (Univerza v Ljubljani, 2005).

27. Yang, H. Feature selection based on joint mutual information. in vol. 23 (1999).

28. Bennasar, M., Hicks, Y. & Setchi, R. Feature selection using Joint Mutual Information Maximisation. Expert Systems with Applications 42, 8520–8532 (2015).

29. N. Spolaôr, E. A. Cherman, M. C. Monard & H. D. Lee. ReliefF for Multi-label Feature Selection. in 6–11 (2013). doi:10.1109/BRACIS.2013.10.

30. Ding, C. & Peng, H. Minimum redundancy feature selection from microarray gene expression data. J Bioinform Comput Biol 3, 185–205 (2005).

31. Xia, S. & Yang, Y. A model-free feature selection technique of feature screening and random forest based recursive feature elimination. (2023).

32. Calzolari, M. Shapicant. (2020).

33. Mayer, M. & Watson, D. kernelshap: Kernel SHAP. (2022).

34. Mayer, M. shapviz: SHAP Visualizations. (2022).

35. Lundberg, S. M. & Lee, S.-I. A unified approach to interpreting model predictions. in Proceedings of the 31st International Conference on Neural Information Processing Systems 4768–4777 (Curran Associates Inc., 2017).

